# Two-Decade Trends and COVID-Era Acceleration in IHD Mortality Among Adults with Alcohol Use Disorder in the United States, 1999–2024

**DOI:** 10.64898/2026.06.22.26356290

**Authors:** Taha Yahya, Syed Ali Raza Zaidi, Suleman Arshad, Meer Hassan Khalid, Mian Zain Hayat, Mohsin Tariq, Muhammad Ahmad, Raghabendra kumar Mahato

**Affiliations:** Shaikh Khalifa Bin Zayed Al-Nahyan Medical College, Shaikh Zayed Postgraduate Medical Institute (SZPGMI), Lahore, Pakistan; Gandaki Medical College Teaching Hospital and Research Center,Pokhara, Nepal

**Keywords:** alcohol use disorder, cardiovascular disease, Centers for Disease Control and Prevention Wide-Ranging Online Data for Epidemiologic Research (CDC WONDER), ischemic heart disease, mortality trends

## Abstract

**Background:** Alcohol use disorder (AUD) is an underrecognized cardiovascular risk factor linked to accelerated atherosclerosis, arrhythmias, and ischemic heart disease (IHD). National trends in IHD mortality among adults with AUD, particularly during the COVID-19 pandemic, remain poorly characterized. We assessed temporal trends and demographic and geographic disparities in IHD-AUD mortality in the United States from 1999 to 2024.

**Methods:** Mortality data for US adults aged ≥25 years were obtained from the CDC WONDER Multiple Cause-of-Death database (1999–2024). Deaths listing both IHD (ICD-10 I20–I25) and AUD (F10) were included. Age-adjusted mortality rates (AAMRs) per 100,000 population were calculated, and Joinpoint regression was used to estimate annual percent changes (APCs) with 95% confidence intervals (CIs).

**Results:** Between 1999 and 2024, 150,273 deaths involved both IHD and AUD. The AAMR declined slightly from 2.0 to 1.9 per 100,000 between 1999 and 2011, increased to 2.7 by 2018, and rose sharply to 3.7 during 2018–2021 (APC, +12.52% [95% CI, 4.97–20.61]; P=0.003), before stabilizing at 3.6 through 2024. Overall mortality increased by approximately 80% from baseline. Mortality increased persistently among adults aged 35–44 years after 2014 (APC, +8.33%) while adults aged 55-64 had the highest mortality rate. Rates were higher in men than women (peak 6.6 vs 1.2 per 100,000). American Indian or Alaska Native individuals had the highest mortality (peak 7.4), whereas Asian or Pacific Islander individuals had the lowest. Black or African American individuals experienced the steepest increase during 2018–2021 (APC, +16.54%). Rates were highest in the West, increased longest in the South, and remained higher in nonmetropolitan than metropolitan areas.

**Conclusion:** IHD mortality among adults with AUD increased substantially over the study period, accelerating during the COVID-19 pandemic. Marked disparities among men, American Indian or Alaska Native and Black or African American individuals, younger adults, and rural populations highlight the need for integrated cardiovascular and addiction care.

## 1 Introduction

Ischemic heart disease (IHD) remains the leading cause of death in the United States, accounting for an estimated 375,476 deaths in 2021 and approximately one in every eight deaths nationwide[1,2]. Although advances in reperfusion strategies and guideline-directed medical therapy have led to substantial reductions in age-adjusted mortality, recent surveillance data suggest that this progress has slowed since 2011, with mortality trends worsening during the COVID-19 pandemic period beginning in 2018 [3,4].

Alcohol use disorder (AUD) affects an estimated 28.9 million adults in the United States and has become an increasingly important contributor to cardiovascular mortality [5,6]. Individuals with AUD face a 1.5-to 2-fold higher risk of IHD through several well-established mechanisms, including myocardial inflammation, mitochondrial dysfunction, hypertension, atrial arrhythmias, and coagulation abnormalities, all of which contribute to atherosclerosis and acute coronary events [7,8]. Growing evidence also highlights a bidirectional relationship between AUD and IHD, with individuals diagnosed with AUD experiencing persistently elevated cardiovascular risk beyond that explained by traditional risk factors alone [9]. This intersection became particularly relevant during the COVID-19 pandemic, when alcohol-related deaths increased by more than 25% between 2019 and 2020 amid major disruptions in addiction services and documented increases in harmful alcohol consumption (Figure 1) [10,11].

**Figure 1.**
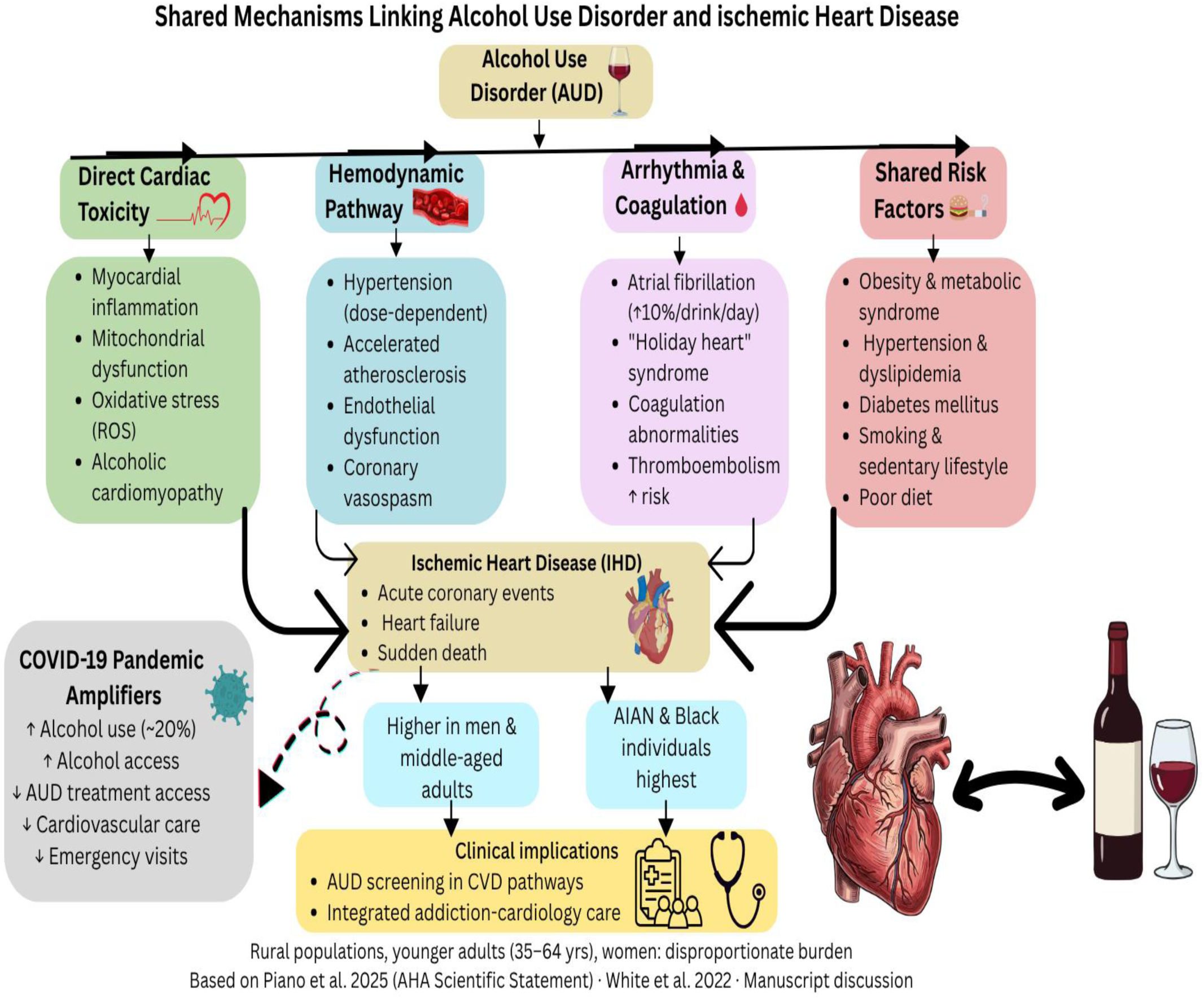
Mechanisms linking alcohol use disorder and ischemic heart disease. AUD indicates alcohol use disorder; and IHD, ischemic heart disease.

Several demographic and geographic factors merit closer examination. Women with AUD appear to experience a disproportionately greater increase in cardiovascular risk than men, contributing to a narrowing of the longstanding sex gap in alcohol-related mortality [12]. Rising IHD mortality among younger adults with AUD is also an emerging concern, particularly given their substantial cumulative alcohol exposure despite a lower perceived cardiovascular risk profile [13]. Marked disparities in both AUD prevalence and IHD outcomes have been reported across racial and ethnic groups, with American Indian and Alaska Native populations experiencing the highest rates of alcohol-attributable mortality [14,15]. Individuals living in rural areas often encounter barriers to both addiction treatment and cardiovascular care, further increasing mortality risk [16]. In addition, geographic variation in AUD-related IHD mortality across census regions and states has important implications for resource allocation and targeted public health interventions [17].

Despite the well-established association between AUD and IHD, no nationwide study has comprehensively evaluated temporal trends in their co-occurring mortality across demographic and geographic subgroups over the full 1999–2024 period, including the COVID-19 pandemic era. Accordingly, we sought to characterize national trends in IHD mortality among adults with AUD listed as a contributing cause of death and to identify disparities by sex, race and ethnicity, age group, census region, state, and urbanization category using Multiple Cause-of-Death data from the Centers for Disease Control and Prevention Wide-Ranging Online Data for Epidemiologic Research (CDC WONDER) [18].

## 2 Methods

### 2.1 Data Source and Study Population

Mortality data were obtained from the CDC WONDER Multiple Cause of Death (MCOD) database for the period 1999–2024. This publicly available dataset contains death certificate records from all 50 US states and the District of Columbia and is compiled by the National Center for Health Statistics (NCHS), which records the underlying cause of death and up to 20 contributing causes for each decedent [18]. Because the data are publicly available and fully de-identified, institutional review board approval was not required. The study was conducted and reported in accordance with the STROBE guidelines [19]. Death records were identified from the MCOD Public Use Files when both ischemic heart disease (IHD; ICD-10 codes I20–I25) and alcohol use disorder (AUD; ICD-10 code F10) were listed anywhere on the death certificate as contributing causes of death, regardless of the underlying cause. This approach captures deaths in which IHD and AUD co-occurred as comorbid conditions and aligns with established MCOD methodologies used in studies of substance use and cardiovascular mortality [20,21]. The study population included US adults aged 25 years or older, consistent with previous cardiovascular mortality analyses using CDC WONDER [20].

### 2.2 Variables and Outcomes

The primary outcome was the annual age-adjusted mortality rate (AAMR) per 100,000 population from 1999 through 2024. Death counts, population denominators, and demographic and geographic variables were extracted from CDC WONDER, standardized across age, sex, and race/ethnicity categories, and exported for analysis. Variables included calendar year; sex; age group (25–34, 35–44, 45–54, 55–64, 65–74, 75–84, and ≥85 years); race/ethnicity (non-Hispanic White, non-Hispanic Black or African American, Hispanic or Latino, non-Hispanic Asian or Pacific Islander, and non-Hispanic American Indian or Alaska Native); urban-rural status according to the 2013 NCHS Urban–Rural Classification Scheme for Counties [22]; US Census region (Northeast, Midwest, South, and West); and state of residence. Crude mortality rates (CMRs) and AAMRs per 100,000 population were calculated for each stratum. AAMRs were derived using direct age standardization to the 2000 US Standard Population [23].

### 2.3 Statistical Analysis

Temporal trends in AAMRs were evaluated using the Joinpoint Regression Program, version 4.9.0.0 (National Cancer Institute, Bethesda, Maryland) [24]. Joinpoint regression fits a series of connected linear segments to the data, with joinpoints representing calendar years in which the direction or magnitude of mortality trends changed significantly. Mortality rates were modeled on the logarithmic scale, which stabilizes variance and permits multiplicative changes to be assessed as linear trends. The slope of each segment was exponentiated and expressed as the annual percent change (APC), representing the estimated yearly change in mortality. Each segment corresponded to a period of statistically homogeneous change until a significant joinpoint was identified. Consistent with National Cancer Institute recommendations, a maximum of three joinpoints was specified for the 26 annual observations spanning 1999–2024. APCs and corresponding 95% confidence intervals (CIs) were estimated using Monte Carlo permutation testing. The average annual percent change (AAPC) and 95% CI were calculated as the weighted average of segment-specific APCs across the study period [25].

In addition to model-derived APC estimates, lag-1 APC values were calculated to characterize short-term year-to-year fluctuations according to:

Lag-1 APC_t_ = [(AAMR_t_ - AAMR_t-1_) / AAMR_t-1_] × 100

where AAMRₜ and AAMR_ₜ₋₁_ represent the age-adjusted mortality rates in year t and the preceding year, respectively. APCs and AAPCs were considered statistically significant when the 95% CI excluded zero. Statistical significance was defined as a two-sided α level of 0.05.

To evaluate the robustness of the primary findings, a prespecified sensitivity analysis was conducted in which AUD was restricted to alcohol dependence syndrome (ICD-10 code F10.2) listed as a contributing cause of death. Consistent with CDC WONDER data-use policies, subgroup estimates based on fewer than 10 deaths in a given year were suppressed. Data visualization was performed using Python version 3.10 (Python Software Foundation).

## 3 Results

### 3.1 Overall Temporal Trends

From 1999 to 2024, a total of 150,273 deaths were attributed to ischemic heart disease with alcohol use disorder as a contributing cause among US adults aged ≥25 years (Table 1;Figure S1,S2). The overall age-adjusted mortality rate (AAMR) demonstrated a triphasic pattern: a non-significant decline from 2.0 per 100,000 in 1999 to 1.9 per 100,000 in 2011, followed by a significant progressive rise through 2018 (AAMR 2.7 per 100,000), and a sharp significant acceleration from 2018 to 2021 (AAMR 3.7 per 100,000; APC: +12.52% [95% CI, 4.97–20.61]; *P*=0.003), before a non-significant plateau through 2024 (AAMR 3.6 per 100,000) representing an approximately 80% net increase from the 1999 baseline (Figure 2;Table S1,S2,S3). Place of death data, available through 2020, revealed that 58.1% of IHD+AUD-related deaths occurred at the decedent’s home, with only 15.0% occurring in an inpatient medical facility (Figure S4)

**Figure 2.**
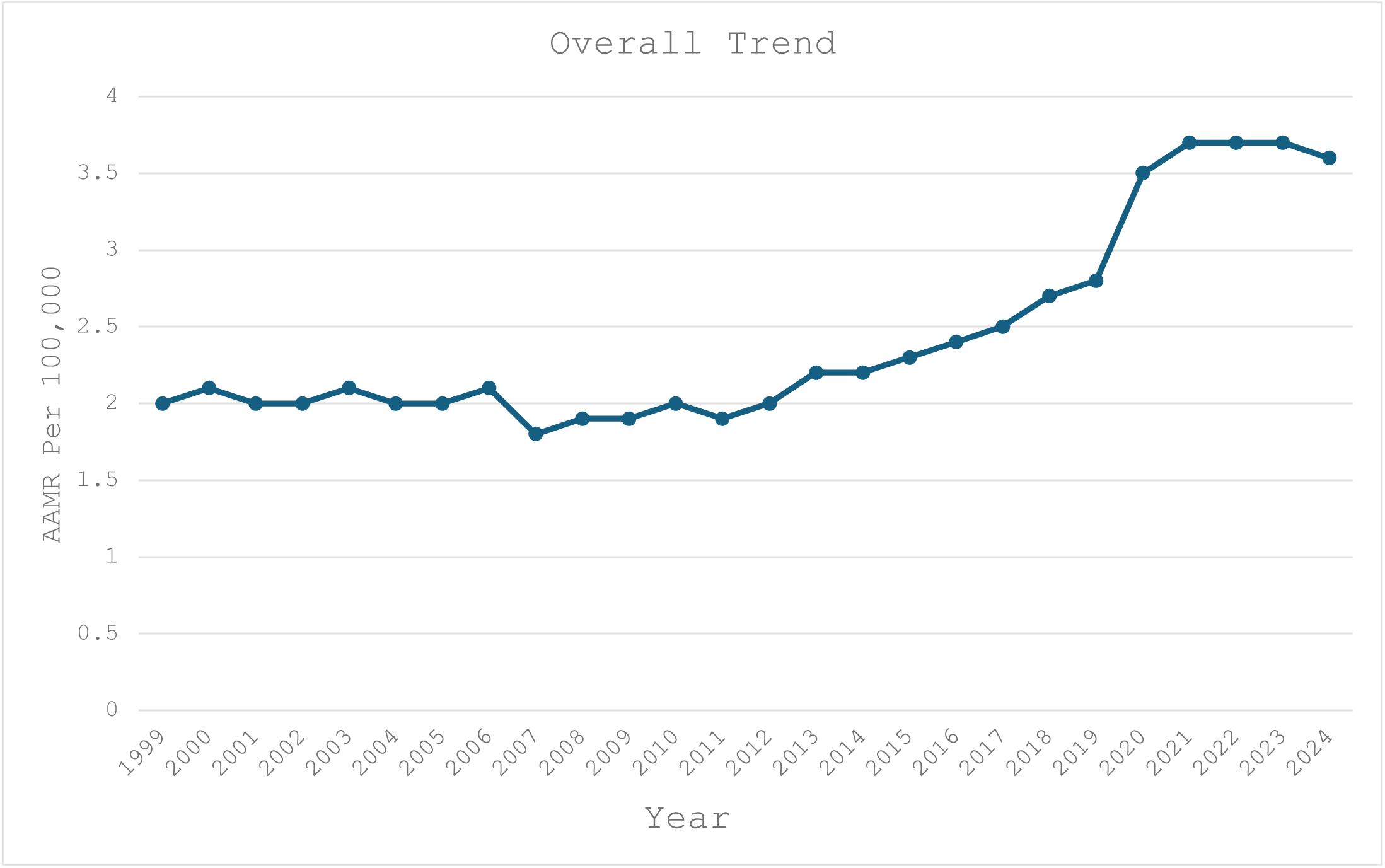
Overall AAMRs per 100,000 and APC (95% CI) for IHD With Alcohol Use Disorder as a Contributing Cause in the United States, 1999 to 2024. Overall: 1999 to 2011 (APC=−0.55 [95% CI, −1.18 to 0.09]), 2011 to 2018 (APC=+4.60* [95% CI, 2.93–6.30]), 2018 to 2021 (APC=+12.52* [95% CI, 4.97–20.61]), and 2021 to 2024 (APC=−1.28 [95% CI, −4.15 to 1.66]). AAMR indicates age-adjusted mortality rate; APC, annual percent change; IHD, ischemic heart disease. *APC significantly different from zero at α=0.05.

**Table 1.**
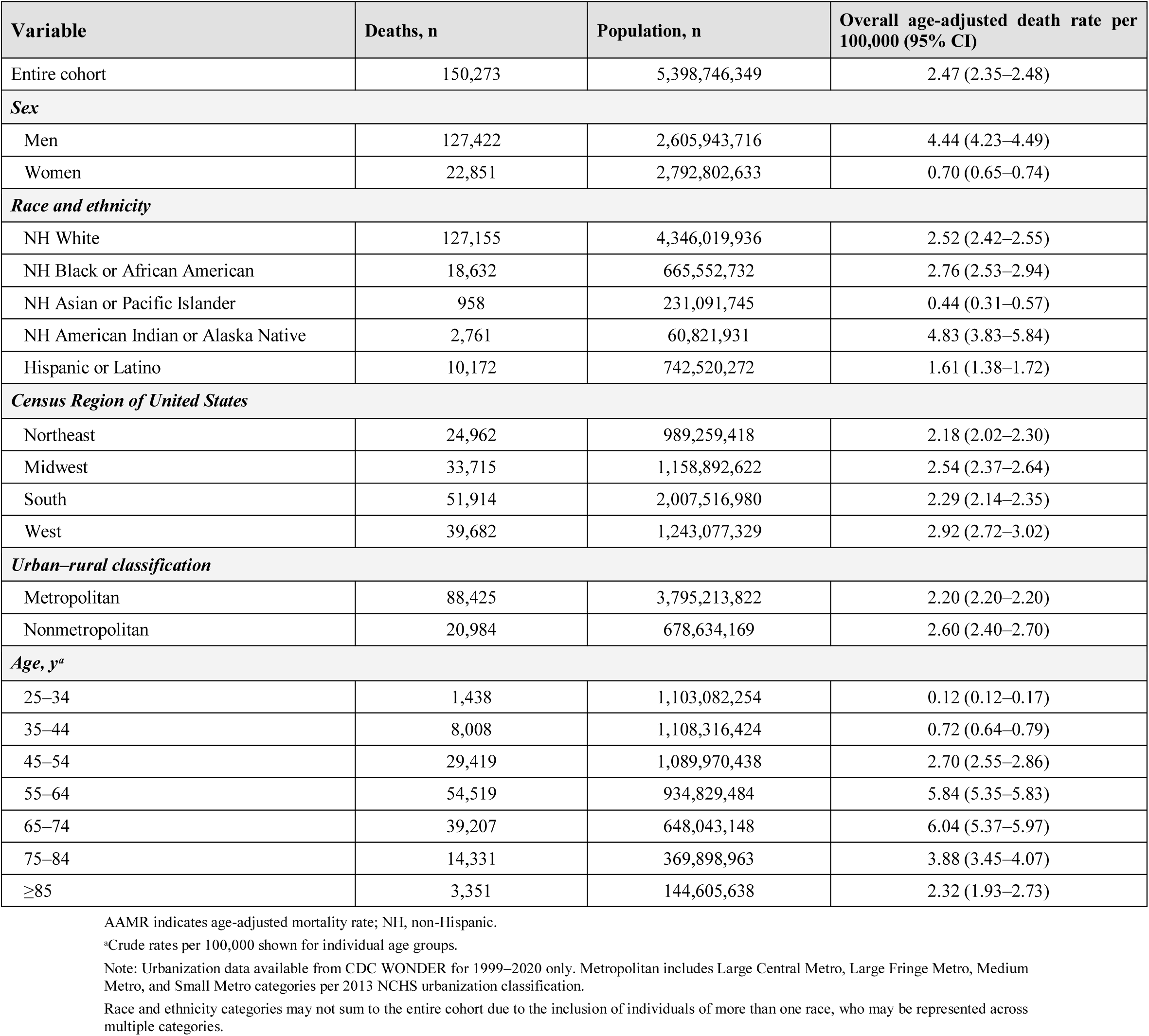
AAMRs per 100,000 individuals and frequency for ischemic heart disease with alcohol-related disorders (ICD-10 codes I20–I25 and F10), United States, 1999–2024.

| Variable | Deaths, n | Population, n | Overall age-adjusted death rate per 100,000 (95% CI) |
| --- | --- | --- | --- |
| Entire cohort | 150,273 | 5,398,746,349 | 2.47 (2.35–2.48) |
| **Sex** |  |  |  |
| Men | 127,422 | 2,605,943,716 | 4.44 (4.23–4.49) |
| Women | 22,851 | 2,792,802,633 | 0.70 (0.65–0.74) |
| **Race and ethnicity** |  |  |  |
| NH White | 127,155 | 4,346,019,936 | 2.52 (2.42–2.55) |
| NH Black or African American | 18,632 | 665,552,732 | 2.76 (2.53–2.94) |
| NH Asian or Pacific Islander | 958 | 231,091,745 | 0.44 (0.31–0.57) |
| NH American Indian or Alaska Native | 2,761 | 60,821,931 | 4.83 (3.83–5.84) |
| Hispanic or Latino | 10,172 | 742,520,272 | 1.61 (1.38–1.72) |
| **Census Region of United States** |  |  |  |
| Northeast | 24,962 | 989,259,418 | 2.18 (2.02–2.30) |
| Midwest | 33,715 | 1,158,892,622 | 2.54 (2.37–2.64) |
| South | 51,914 | 2,007,516,980 | 2.29 (2.14–2.35) |
| West | 39,682 | 1,243,077,329 | 2.92 (2.72–3.02) |
| **Urban–rural classification** |  |  |  |
| Metropolitan | 88,425 | 3,795,213,822 | 2.20 (2.20–2.20) |
| Nonmetropolitan | 20,984 | 678,634,169 | 2.60 (2.40–2.70) |
| **Age, y<sup>a</sup>** |  |  |  |
| 25–34 | 1,438 | 1,103,082,254 | 0.12 (0.12–0.17) |
| 35–44 | 8,008 | 1,108,316,424 | 0.72 (0.64–0.79) |
| 45–54 | 29,419 | 1,089,970,438 | 2.70 (2.55–2.86) |
| 55–64 | 54,519 | 934,829,484 | 5.84 (5.35–5.83) |
| 65–74 | 39,207 | 648,043,148 | 6.04 (5.37–5.97) |
| 75–84 | 14,331 | 369,898,963 | 3.88 (3.45–4.07) |
| ≥85 | 3,351 | 144,605,638 | 2.32 (1.93–2.73) |

AAMR indicates age-adjusted mortality rate; NH, non-Hispanic.
<sup>a</sup>Crude rates per 100,000 shown for individual age groups.
Note: Urbanization data available from CDC WONDER for 1999–2020 only. Metropolitan includes Large Central Metro, Large Fringe Metro, Medium Metro, and Small Metro categories per 2013 NCHS urbanization classification.
Race and ethnicity categories may not sum to the entire cohort due to the inclusion of individuals of more than one race, who may be represented across multiple categories.

### 3.2 Age Group-Stratified Trends

Crude mortality rates were heterogeneous across all seven age groups over the study period. Adults aged 55–64 years carried the highest absolute death burden among working-age groups (n=54,519) and demonstrated the most sustained acceleration in mortality, with rates rising significantly from 2011 to 2022 (APC=+7.47% [95% CI, 6.31–8.64]). Adults aged 35–44 years exhibited a notable reversal from an initial declining trajectory, with a sharp and significant rise from 2014 through 2024 (APC=+8.33% [95% CI, 6.50–10.19]). Adults aged 45–54 years showed a sustained uninterrupted increase from 1999 to 2018 (APC=+1.21% [95% CI, 0.77–1.66]), together with the 55–64 and 35–44 age groups underscoring a broad and worsening middle-age burden. In contrast, adults aged ≥85 years were the only group to exhibit an initial significant decline from 1999 to 2008 (APC=−8.40% [95% CI, −12.76 to −3.82]) before reversing to a significant rise through 2024 (APC=+5.14% [95% CI, 3.25–7.06]), representing the most paradoxical trajectory across all age groups. Adults aged 65–74 and 75–84 years also demonstrated significant increases in more recent periods, while adults aged 25–34 years showed the lowest absolute rates throughout the study period (Figure 3;Table S2,S4).

**Figure 3.**
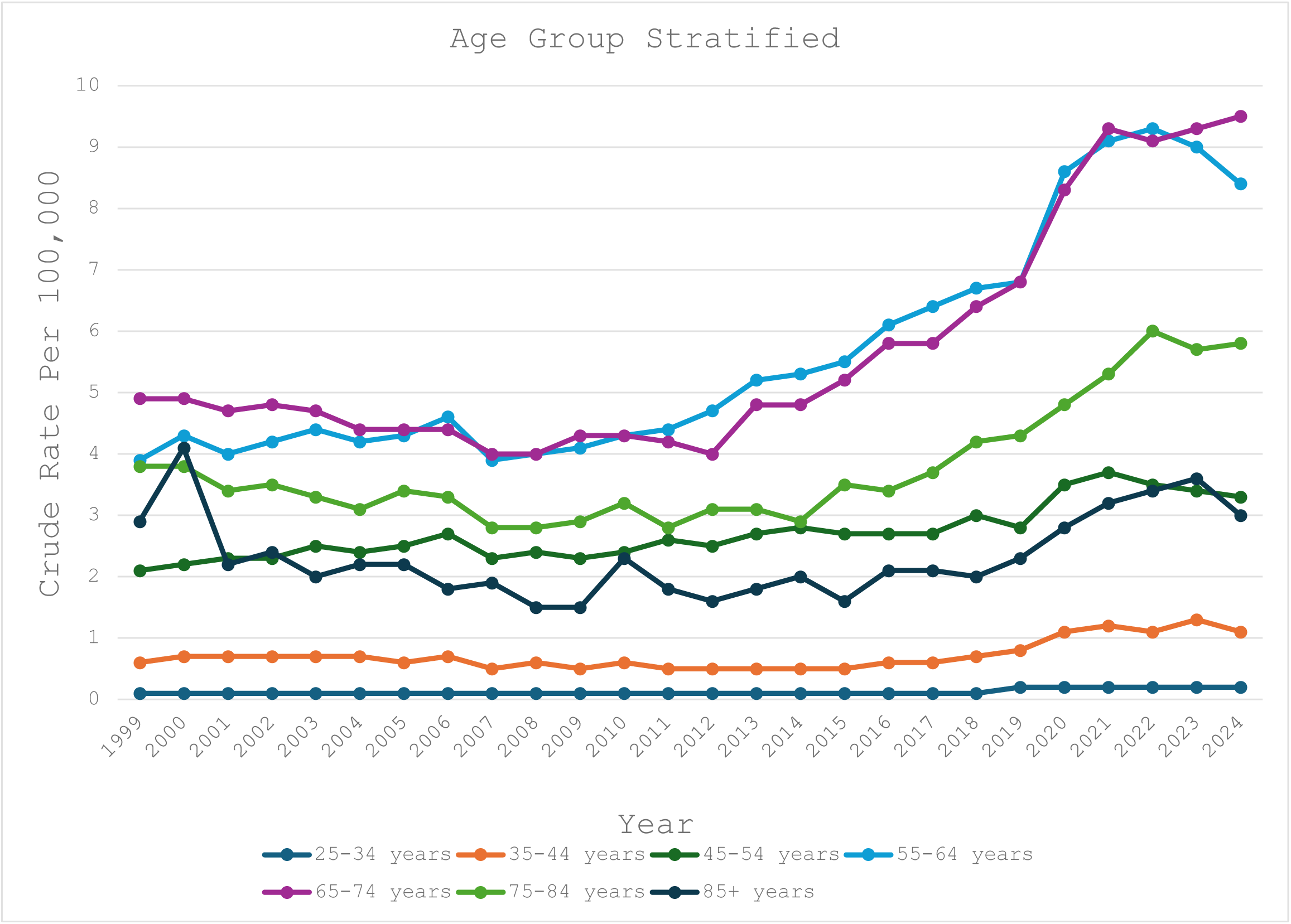
Crude Mortality Rates per 100,000 and APC (95% CI) Stratified by Age Group for IHD With Alcohol Use Disorder as a Contributing Cause in the United States, 1999 to 2024. Age 25 to 34 years: 1999 to 2017 (APC=+0.12* [95% CI, 0.12–0.12]), 2017 to 2020 (APC=+27.98* [95% CI, 27.98–27.98]), and 2020 to 2024 (APC=−2.02* [95% CI, −2.02 to −2.02]). Age 35 to 44 years: 1999 to 2014 (APC=−2.44* [95% CI, −3.47 to −1.39]) and 2014 to 2024 (APC=+8.33* [95% CI, 6.50–10.19]). Age 45 to 54 years: 1999 to 2018 (APC=+1.21* [95% CI, 0.77–1.66]). Age 55 to 64 years: 2011 to 2022 (APC=+7.47* [95% CI, 6.31–8.64]). Age 65 to 74 years: 1999 to 2011 (APC=−1.43* [95% CI, −2.37 to −0.47]), 2011 to 2018 (APC=+6.42* [95% CI, 4.60–8.28]), and 2018 to 2021 (APC=+13.10* [95% CI, 2.29–25.05]). Age 75 to 84 years: 1999 to 2008 (APC=−3.23* [95% CI, −4.50 to −1.94]) and 2014 to 2022 (APC=+8.75* [95% CI, 6.67–10.88]). Age ≥85 years: 1999 to 2008 (APC=−8.40* [95% CI, −12.76 to −3.82]) and 2008 to 2024 (APC=+5.14* [95% CI, 3.25–7.06]). APC indicates annual percent change; IHD, ischemic heart disease. *APC significantly different from zero at α=0.05.

### 3.3 Sex-Stratified Trends

Males demonstrated higher AAMR than females throughout the study period, with the absolute sex disparity widening substantially over time. Among males, who carried the higher burden (AAMR 6.6 per 100,000 at peak), a significant acceleration was observed from 2018 to 2021 (APC: +12.24% [95% CI, 2.71–22.65]; *P*=0.01), with the AAMR remaining elevated at 6.3 per 100,000 by 2024. Among females, the AAMR remained stable from 1999 to 2012 before a steep significant rise through 2022 (APC: +9.45% [95% CI, 7.47–11.48]; *P*<0.000001), reaching 1.2 per 100,000 (Figure 4;Table S1,S2,S3).

**Figure 4.**
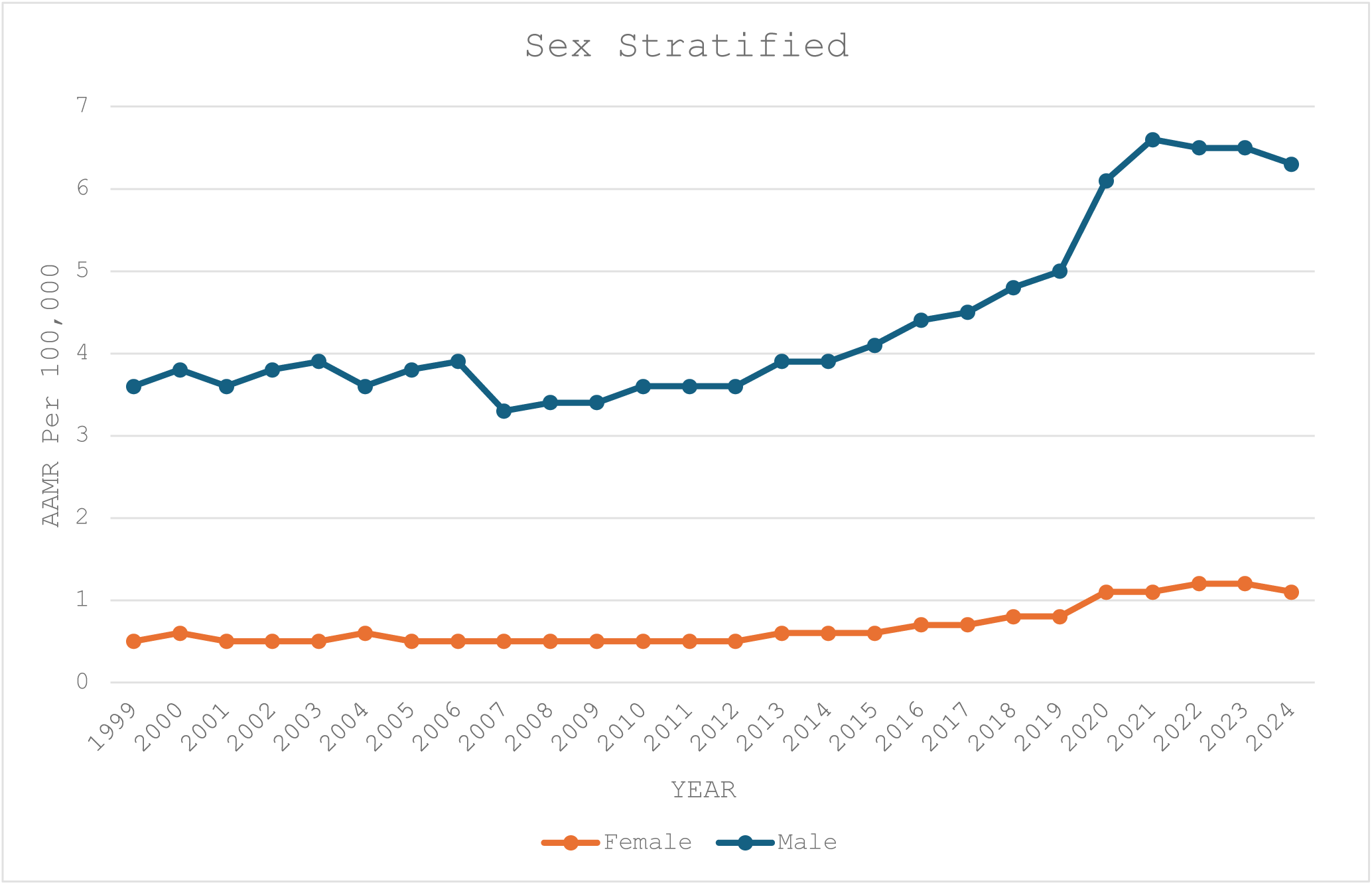
AAMRs per 100,000 and APC (95% CI) Stratified by Sex for IHD With Alcohol Use Disorder as a Contributing Cause in the United States, 1999 to 2024. Male: 1999 to 2011 (APC=−0.61 [95% CI, −1.40 to 0.19]), 2011 to 2018 (APC=+4.23* [95% CI, 2.14–6.36]), 2018 to 2021 (APC=+12.24* [95% CI, 2.71–22.65]), and 2021 to 2024 (APC=−1.58 [95% CI, −5.24 to 2.22]). Female: 1999 to 2012 (APC=−0.84 [95% CI, −2.41 to 0.76]), 2012 to 2022 (APC=+9.45* [95% CI, 7.47–11.48]), and 2022 to 2024 (APC=−3.53 [95% CI, −15.86 to 10.61]). AAMR indicates age-adjusted mortality rate; APC, annual percent change; IHD, ischemic heart disease. *APC significantly different from zero at α=0.05.

### 3.4 Race/Ethnicity-Stratified Trends

American Indian or Alaska Native individuals carried the highest AAMR of all racial and ethnic groups throughout the study period, peaking at 7.7 per 100,000 in 2023, while Asian or Pacific Islander individuals demonstrated the lowest rates (0.3–0.8 per 100,000). Black or African American individuals experienced the sharpest single-interval acceleration, with a surge of +16.54% annually (95% CI, 0.75–34.82; *P*=0.04) between 2018 and 2021, reaching 3.9 per 100,000. Hispanic or Latino individuals demonstrated the steepest consistent rise of any group from 2014 to 2022 (APC: +10.82% [95% CI, 7.94–13.77]; *P*<0.000001), reaching 2.5 per 100,000 (Figure 5;Table S1,S2,S3).

**Figure 5.**
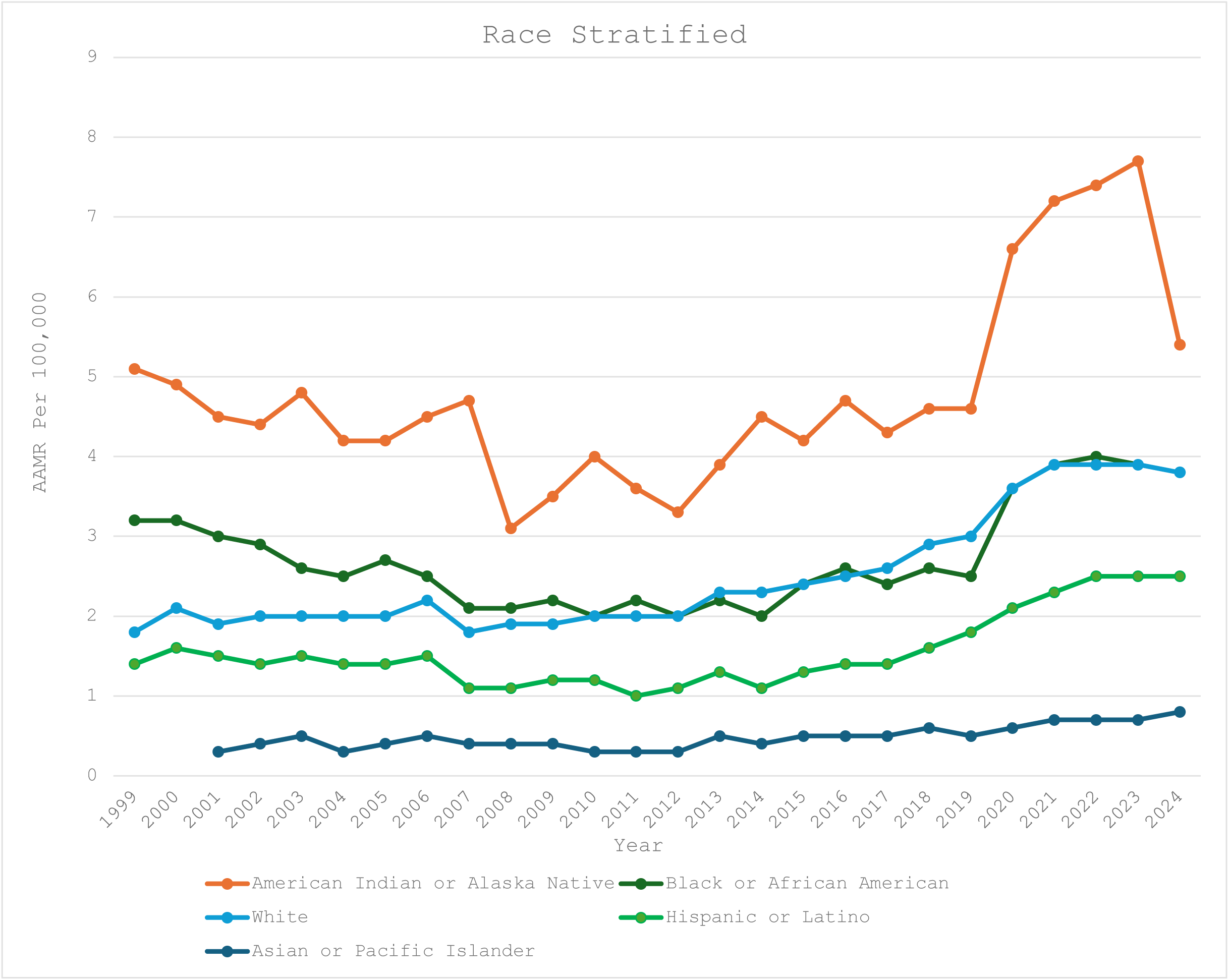
AAMRs per 100,000 and APC (95% CI) Stratified by Race and Ethnicity for IHD With Alcohol Use Disorder as a Contributing Cause in the United States, 1999 to 2024. American Indian or Alaska Native: 1999 to 2011 (APC=−2.73* [95% CI, −4.64 to −0.78]), 2011 to 2019 (APC=+4.25* [95% CI, 0.83–7.79]), and 2022 to 2024 (APC=−16.12* [95% CI, −29.58 to −0.09]). Asian or Pacific Islander: 2001 to 2020 (APC=+2.09* [95% CI, 0.72–3.48]). Black or African American: 1999 to 2010 (APC=−4.27* [95% CI, −5.31 to −3.21]), 2010 to 2018 (APC=+2.80* [95% CI, 0.29–5.37]), and 2018 to 2021 (APC=+16.54* [95% CI, 0.75–34.82]). White: 2011 to 2018 (APC=+5.04* [95% CI, 2.60–7.54]) and 2018 to 2021 (APC=+12.07* [95% CI, 1.52–23.72]). Hispanic or Latino: 1999 to 2014 (APC=−2.24* [95% CI, −3.28 to −1.18]) and 2014 to 2022 (APC=+10.82* [95% CI, 7.94–13.77]). AAMR indicates age-adjusted mortality rate; APC, annual percent change; IHD, ischemic heart disease. *APC significantly different from zero at α=0.05

### 3.5 Census Region-Stratified Trends

The West sustained the highest AAMR among all census regions throughout the study period, peaking at 4.3 per 100,000 in 2021 with a sharp significant acceleration from 2018 to 2021 (APC: +12.35% [95% CI, 0.59–25.49]; *P*=0.04). The South demonstrated the most prolonged uninterrupted rise of any region, from 2011 through 2024 (APC: +3.72% [95% CI, 3.28–4.17]; *P*<0.000001), while the Midwest exhibited the highest single-period acceleration (APC: +11.80% [95% CI, −0.23 to 25.29] from 2018 to 2021), which did not reach statistical significance (*P*=0.054) (Figure 6;Table S2,S7).

**Figure 6.**
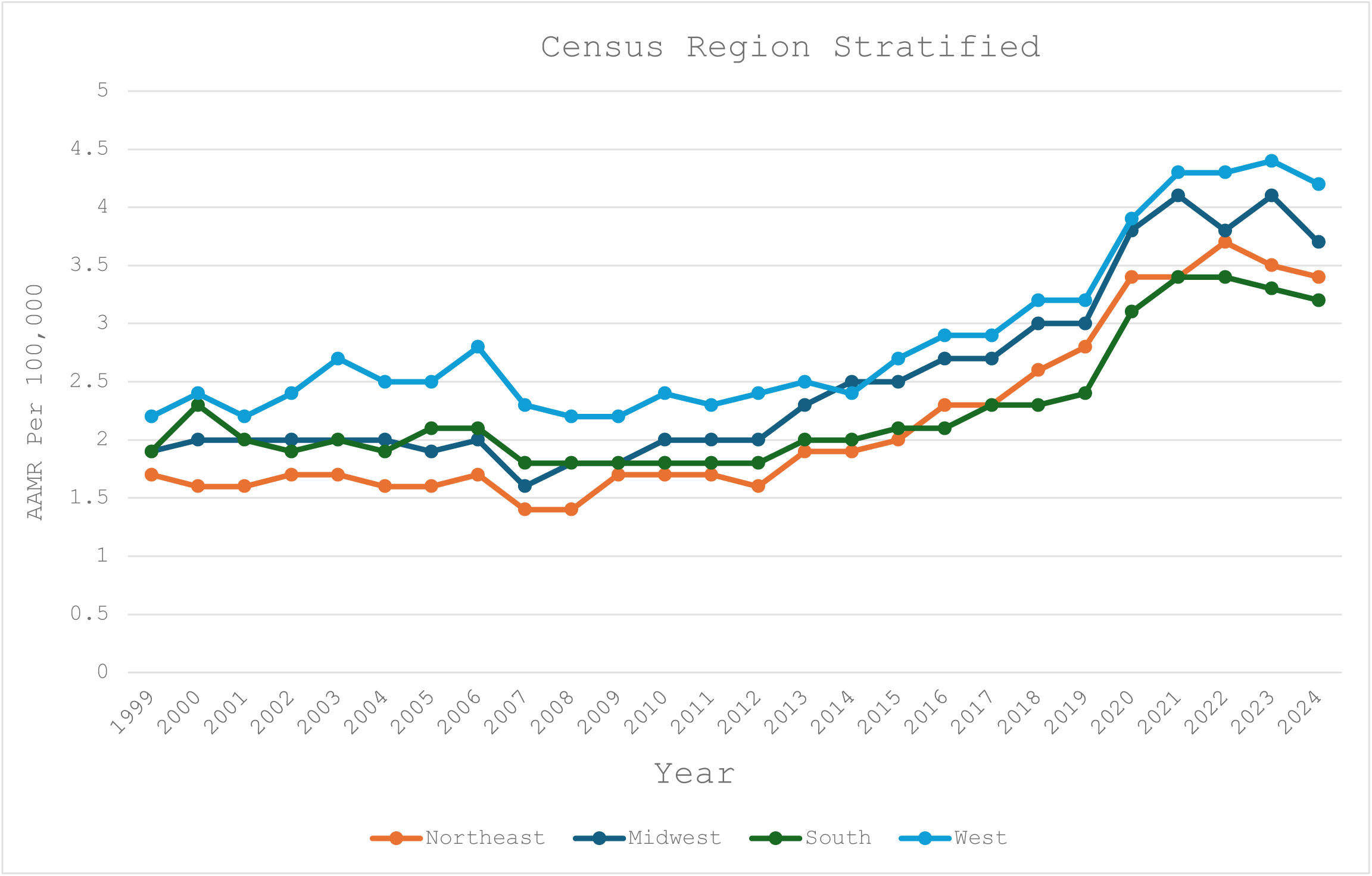
AAMRs per 100,000 and APC (95% CI) Stratified by Census Region for IHD With Alcohol Use Disorder as a Contributing Cause in the United States, 1999 to 2024. Northeast: 1999 to 2012 (APC=−0.11 [95% CI, −1.23 to 1.03]), 2012 to 2022 (APC=+8.68* [95% CI, 7.25–10.12]), and 2022 to 2024 (APC=−4.76 [95% CI, −14.14 to 5.65]). Midwest: 1999 to 2009 (APC=−0.98 [95% CI, −2.54 to 0.60]), 2009 to 2018 (APC=+5.35* [95% CI, 3.47–7.26]), 2018 to 2021 (APC=+11.80 [95% CI, −0.23 to 25.29]), and 2021 to 2024 (APC=−2.42 [95% CI, −7.23 to 2.64]). South: 1999 to 2011 (APC=−0.04 [95% CI, −1.49 to 1.43]) and 2011 to 2024 (APC=+3.72* [95% CI, 3.28–4.17]). West: 1999 to 2006 (APC=+2.46* [95% CI, 0.25–4.71]), 2006 to 2009 (APC=−7.05 [95% CI, −21.75 to 10.40]), 2009 to 2018 (APC=+3.98* [95% CI, 2.31–5.68]), 2018 to 2021 (APC=+12.35* [95% CI, 0.59–25.49]), and 2021 to 2024 (APC=−0.31 [95% CI, −4.75 to 4.33]). AAMR indicates age-adjusted mortality rate; APC, annual percent change; IHD, ischemic heart disease. *APC significantly different from zero at α=0.05.

### 3.6 Urbanization-Stratified Trends

Non-metropolitan areas consistently bore a higher AAMR than metropolitan areas throughout the study period. Non-metropolitan communities showed an earlier inflection point (2011; APC: +3.92% [95% CI, 1.06–6.87]; *P*=0.011) compared with metropolitan areas (2012; APC: +6.55%

[95% CI, 5.07–8.05]; *P*<0.000001), and demonstrated the sharper final increase (2018–2020; APC: +17.54% [95% CI, 4.61–32.07]; *P*=0.011), reaching 4.2 per 100,000 by 2020 — 27% higher than the metropolitan AAMR of 3.3 per 100,000 at the same timepoint (Figure 7;Table S2,S5).

**Figure 7.**
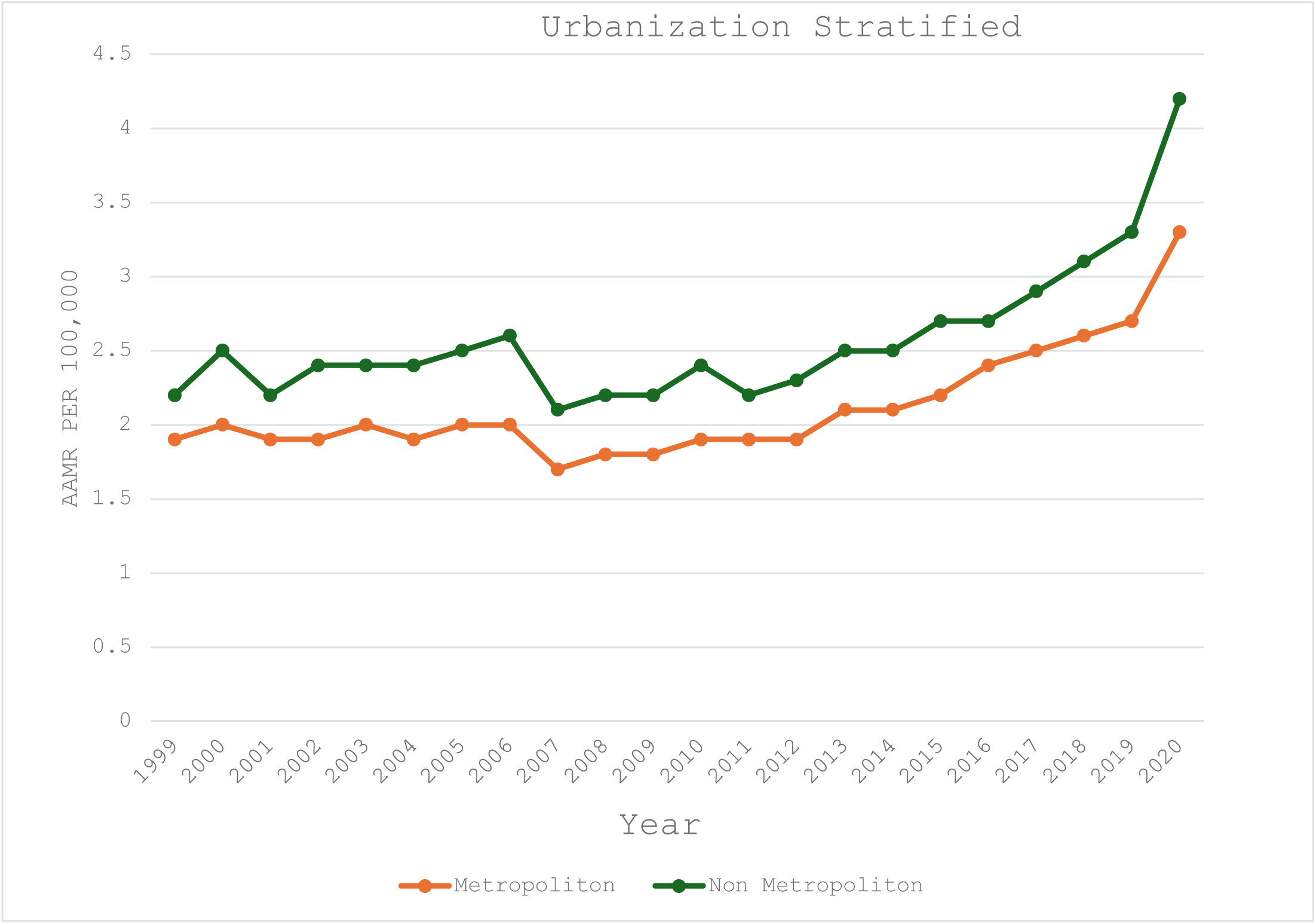
AAMRs per 100,000 and APC (95% CI) Stratified by Urban–Rural Classification for IHD With Alcohol Use Disorder as a Contributing Cause in the United States, 1999 to 2020. Metropolitan: 1999 to 2012 (APC=−0.39 [95% CI, −1.28 to 0.50]) and 2012 to 2020 (APC=+6.55* [95% CI, 5.07–8.05]). Nonmetropolitan: 1999 to 2011 (APC=−0.43 [95% CI, −1.52 to 0.66]), 2011 to 2018 (APC=+3.92* [95% CI, 1.06–6.87]), and 2018 to 2020 (APC=+17.54* [95% CI, 4.61–32.07]). AAMR indicates age-adjusted mortality rate; APC, annual percent change; IHD, ischemic heart disease. *APC significantly different from zero at α=0.05.

### 3.7 State-Level Trends

Among all 50 states and the District of Columbia, the AAMR varied more than 5-fold, from 0.9 per 100,000 in Alabama to 5.2 per 100,000 in Vermont. States in the highest mortality tier (≥90th percentile) included Vermont, North Dakota, Alaska, the District of Columbia, Montana, Nevada, and New Mexico; those in the lowest tier (≤10th percentile) included Alabama, Hawaii, Nebraska, Georgia, Illinois, New Jersey, and Pennsylvania. California recorded the highest absolute death count (n=12,408; 11.3% of total deaths), followed by Florida (n=7,040; 6.4%), New York (n=6,924; 6.3%), Texas (n=6,246; 5.7%), and Ohio (n=4,508; 4.1%) (Figure 8;Table S6).

**Figure 8.**
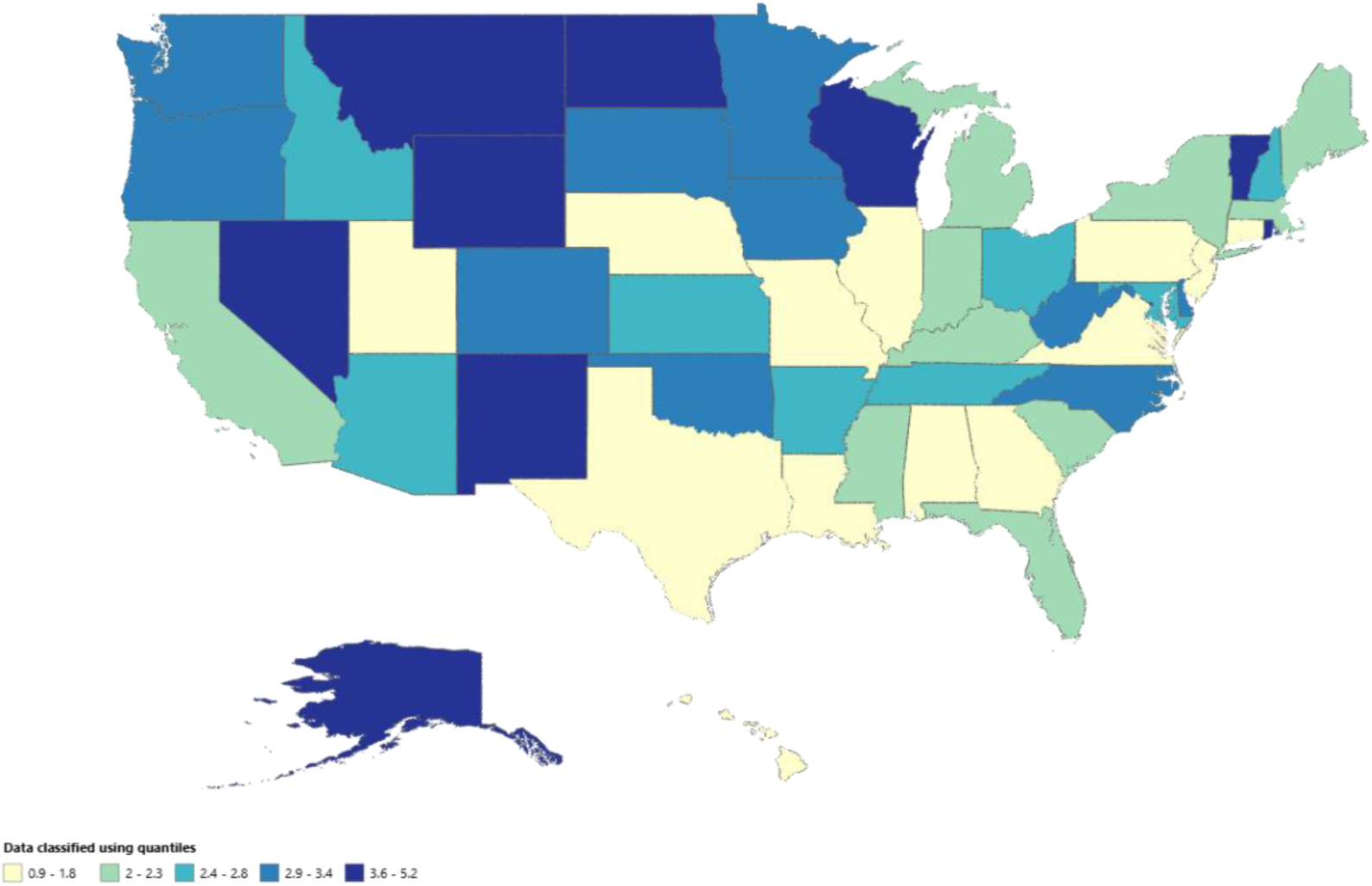
State-Level IHD With Alcohol Use Disorder AAMRs per 100,000 People (Range: 0.9–5.2 per 100,000 People) in the United States, 1999 to 2020. Lighter shading signifies a lower AAMR and darker shading signifies a higher AAMR. AAMR indicates age-adjusted mortality rate; IHD, ischemic heart disease.

## 4 Discussion

In this comprehensive nationwide analysis (Figure 9), we identified 150,273 deaths among US adults in which ischemic heart disease (IHD) and alcohol use disorder (AUD) co-occurred from 1999 through 2024, representing an approximately 80% net increase in age-adjusted mortality from the 1999 baseline. These findings contrast sharply with the broader decline in cardiovascular mortality reported in the 2024 American Heart Association Heart Disease and Stroke Statistics update [26]. To our knowledge, this is the first nationwide study to characterize long-term IHD mortality trends in the presence of AUD across a 26-year period that includes the COVID-19 pandemic and suggests that AUD represents an increasingly important and underrecognized modifiable contributor to cardiovascular mortality in the United States. Although Minhas et al previously reported a 4% annual increase in substance use–related cardiovascular mortality between 1999 and 2019 despite declining overall cardiovascular disease mortality [20], our findings extend those observations by incorporating a longer follow-up period and pandemic-era trends.

**Figure 9.**
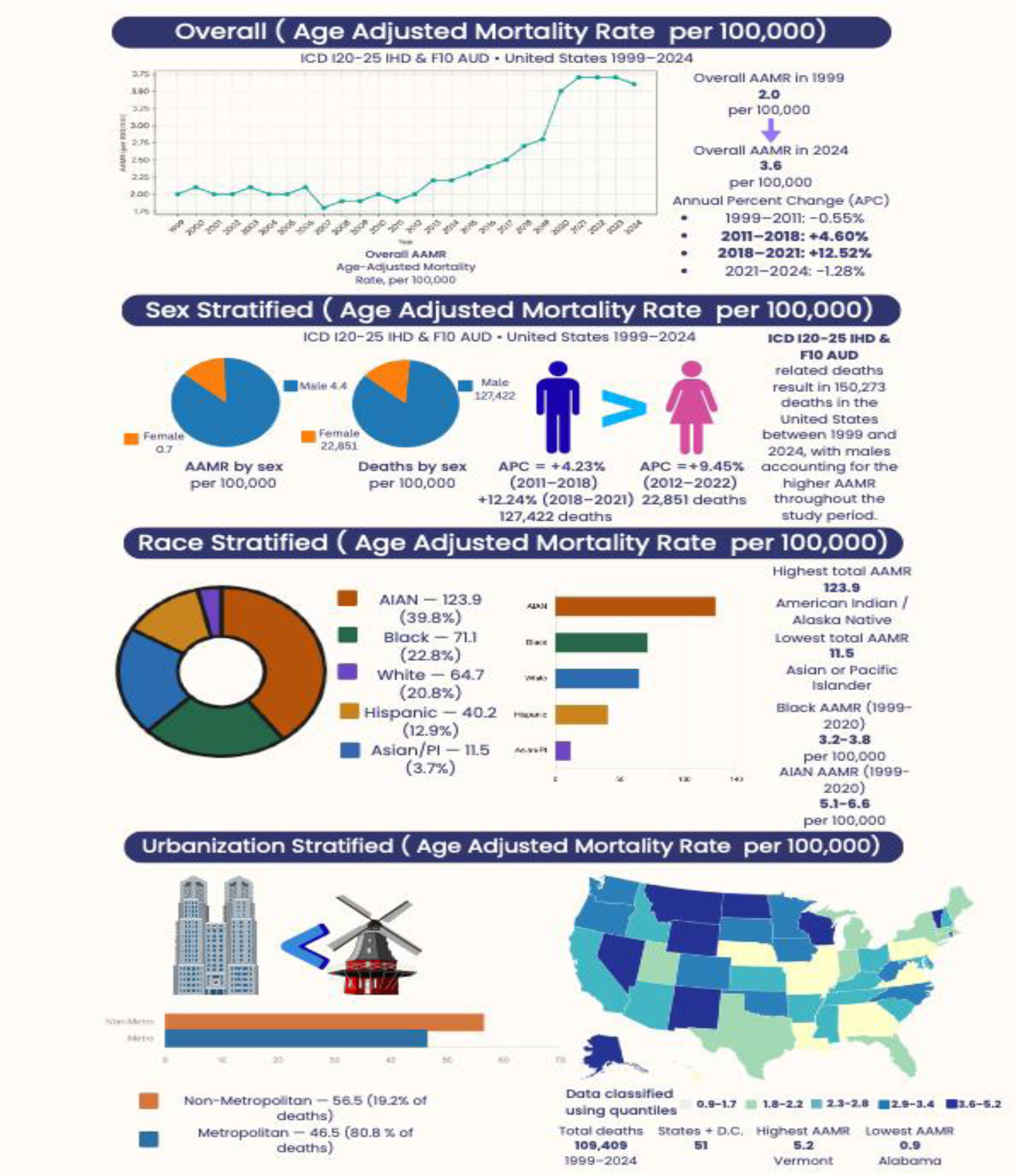
Central illustration summarizing the multi-decadal epidemiological landscape, demographic disparities, and geographic distribution of ischemic heart disease mortality with co-occurring alcohol use disorder in the United States, 1999–2024.

The most notable finding was the sharp acceleration in IHD mortality associated with AUD between 2018 and 2021, coinciding with the COVID-19 pandemic. White et al reported a 25.5% increase in alcohol-related deaths between 2019 and 2020 alone, nearly 12 times the average annual increase observed before the pandemic, with adults aged 35–44 years experiencing the largest rise of any demographic group [27]. Similarly, the Centers for Disease Control and Prevention reported an approximately 23% increase in deaths attributable to excessive alcohol use between 2018–2019 and 2020–2021, roughly four times the increase observed during the preceding biennium [28]. Several factors likely contributed to this surge. According to the 2025 American Heart Association scientific statement by Piano et al, heavy alcohol consumption is strongly associated with hypertension, arrhythmias, coronary vasospasm, alcoholic cardiomyopathy, accelerated atherosclerosis, and other cardiovascular complications, while heavy alcohol use increased by approximately 20% during the pandemic [29]. At the same time, liquor retailers were classified as essential businesses in most states and alcohol delivery services expanded substantially, reducing barriers to alcohol access among individuals with AUD [30]. Access to AUD treatment also declined during the pandemic despite increased telehealth availability [31], while delayed cardiovascular evaluation and avoidance of emergency care further increased cardiovascular risk, particularly in rural communities [16]. The absence of a significant decline after 2021 emphasizes that the excess mortality burden accumulated during the pandemic has persisted rather than returning to pre-pandemic trajectories. Additionally, the predominance of home deaths further illustrates that meaningful reduction in IHD+AUD mortality will require upstream prevention and outpatient AUD screening rather than acute hospital based intervention alone.

The concentration of mortality burden in middle-aged adults particularly those aged 35–64 years represents one of the most clinically significant findings of this analysis. The persistent increase among adults aged 55–64 years, predating the pandemic by nearly a decade, suggests that structural and behavioral factors independent of COVID-19 have been driving excess IHD mortality in this cohort for years in working-age US adults. This prolonged rise likely reflects the cumulative cardiovascular effects of long-term heavy alcohol exposure superimposed on worsening cardiometabolic risk. Consistent with this interpretation, Piano et al reported that chronic alcohol consumption exceeding 80 g/day for at least five years contributes to alcoholic cardiomyopathy and a dilated cardiomyopathy phenotype characterized by impaired myocardial contractility [29]. Adults aged 35–44 years exhibited a transition from declining mortality to a significant upward trajectory beginning in 2014 and experienced the steepest proportional increase during the pandemic. White et al [11] similarly identified this age group as having the largest increase in alcohol-related mortality nationally during 2020, suggesting that pandemic-related changes in alcohol consumption and disruptions in healthcare access disproportionately affected younger adults with AUD [27]. Adults aged 45–54 years also demonstrated a persistent upward trend throughout the study period, without evidence of plateauing. Collectively, these findings identify adults aged 35–64 years as the subgroup experiencing the most dynamic and clinically concerning increases in AUD-associated IHD mortality. Conversely, the oldest age group (≥85 years) was the only subgroup to show an early improvement before subsequently worsening, highlighting the growing concentration of AUD-related IHD mortality among middle-aged rather than oldest-old adults. These findings challenge traditional risk frameworks that primarily associate IHD screening and prevention efforts with older populations. Similar observations were reported by Jain et al, who identified middle-aged adults as an increasingly vulnerable subgroup in national cardiovascular mortality analyses from 1999 to 2019 [32].

Sex-stratified analyses showed that men consistently experienced higher absolute mortality rates; however, women demonstrated a steeper proportional increase, with significant acceleration from 2012 through 2022. This diminishing sex disparity accords with the telescoping phenomenon described by Towers et al, whereby women progress more rapidly from alcohol initiation to dependence and alcohol-related organ injury than men, often developing comparable or greater cardiovascular damage despite lower cumulative alcohol exposure [33]. These findings also raise the possibility that women with AUD remain underrecognized in cardiovascular risk assessment because IHD continues to be viewed predominantly through a male-centered clinical framework.

Substantial racial and ethnic disparities were observed throughout the study period. American Indian or Alaska Native individuals recorded the highest mortality rates of all racial and ethnic groups, with rates peaking in 2022. Karaye et al reported that this population was 3.6 times more likely to die from alcohol-related causes than non-Hispanic White individuals between 1999 and 2020, a disparity linked to structural barriers including poverty, geographic isolation, and limited access to addiction treatment and cardiovascular care [34]. Black or African American individuals exhibited the steepest increase during the 2018–2021 interval, aligning with reports of disproportionate pandemic-era increases in alcohol-related mortality [27]. Hispanic or Latino individuals demonstrated the most sustained increase from 2014 through 2022, indicating that the burden extends across multiple populations rather than being concentrated within a single racial or ethnic group.

Geographic analyses revealed persistently higher mortality rates in nonmetropolitan areas, accompanied by a more pronounced increase during the pandemic. Rural–urban disparities in cardiovascular mortality have widened steadily since 1999 [16], and our findings emphasize that AUD may be an important contributor to this divergence. Limited access to specialty cardiovascular services and evidence-based AUD treatment in rural communities likely further amplifies this risk [35].

### 4.1 Limitations

Several limitations should be considered. First, CDC WONDER relies on death certificate data, which are subject to inaccuracies and incomplete reporting. AUD is known to be underreported on death certificates, potentially leading to underestimation of the true mortality burden. Misclassification of ICD-10 codes may also introduce bias. Second, important clinical variables including AUD severity, treatment history, IHD subtype, medication use, and revascularization status are not available within CDC WONDER and therefore could not be incorporated into the analysis. Third, because this study used an ecological design, causal inferences at the individual level cannot be made. Finally, urban-rural analyses were limited to data available through 2020, which may not fully capture geographic patterns during the later post-pandemic period.

### 4.2 Future Directions

These findings have important implications for both clinical practice and public health policy. Routine screening for AUD within cardiovascular care pathways may be particularly valuable for adults aged 35–64 years, women, American Indian or Alaska Native individuals, and rural populations. The observed associations support recognition of AUD as a clinically relevant and potentially modifiable cardiovascular risk factor that warrants systematic assessment alongside established risk factors such as hypertension and dyslipidemia [29]. The persistence of elevated mortality following the pandemic peak further affirms the need to expand access to evidence based AUD treatment, particularly in rural and underserved communities. In addition, the temporal association between increasing mortality and expanded alcohol availability during the pandemic warrants further evaluation of alcohol delivery and access policies that may have contributed to these trends [28,31].

## 5 Conclusion

IHD mortality among US adults with co-occurring AUD increased by approximately 80% between 1999 and 2024, with the most pronounced acceleration occurring during the COVID-19 pandemic. The burden was concentrated among middle-aged adults and disproportionately affected women, American Indian or Alaska Native individuals, and residents of rural communities. These findings highlight the growing intersection between addiction and cardiovascular disease and support greater integration of cardiovascular and addiction care, alongside targeted prevention and treatment strategies for populations at highest risk.

## ACKNOWLEDGEMENTS

None

## AUTHOR CONTRIBUTIONS

Author Taha Yahya contributed to Writing-Original Draft, Writing-Review and Editing, Conceptualization, Software Analysis, Visualization, Validation, Supervision, and Data Curation. Author Syed Ali Raza Zaidi contributed to Project Administration, Writing-Original Draft, Writing-Review and Editing, Conceptualization, Software Analysis, and Visualization. Author Suleman Arshad contributed to Writing-Original Draft, Conceptualization, Visualization, Validation, and Investigation. Author Meer Hassan Khalid contributed to Data Curation, Software Analysis, Methodology, and Visualization. Author Mian Zain Hayat contributed to Data Curation, and Resources. Author Mohsin Tariq contributed to Investigation, and Resources. Author Muhammad Ahmad contributed to Methodology and Visualization.

Author Raghabendra Kumar Mahato contributed to Software Analysis and Visualization.

## DATA AVAILABILITY STATEMENT

Data sharing does not apply to this article as no datasets were generated during the current study; the data that support the findings of this study are publicly available from the CDC WONDER Multiple Cause-of-Death database. Analytic code and non-identifiable derived data are available from the corresponding author upon reasonable request.

---

**Figure S1:**
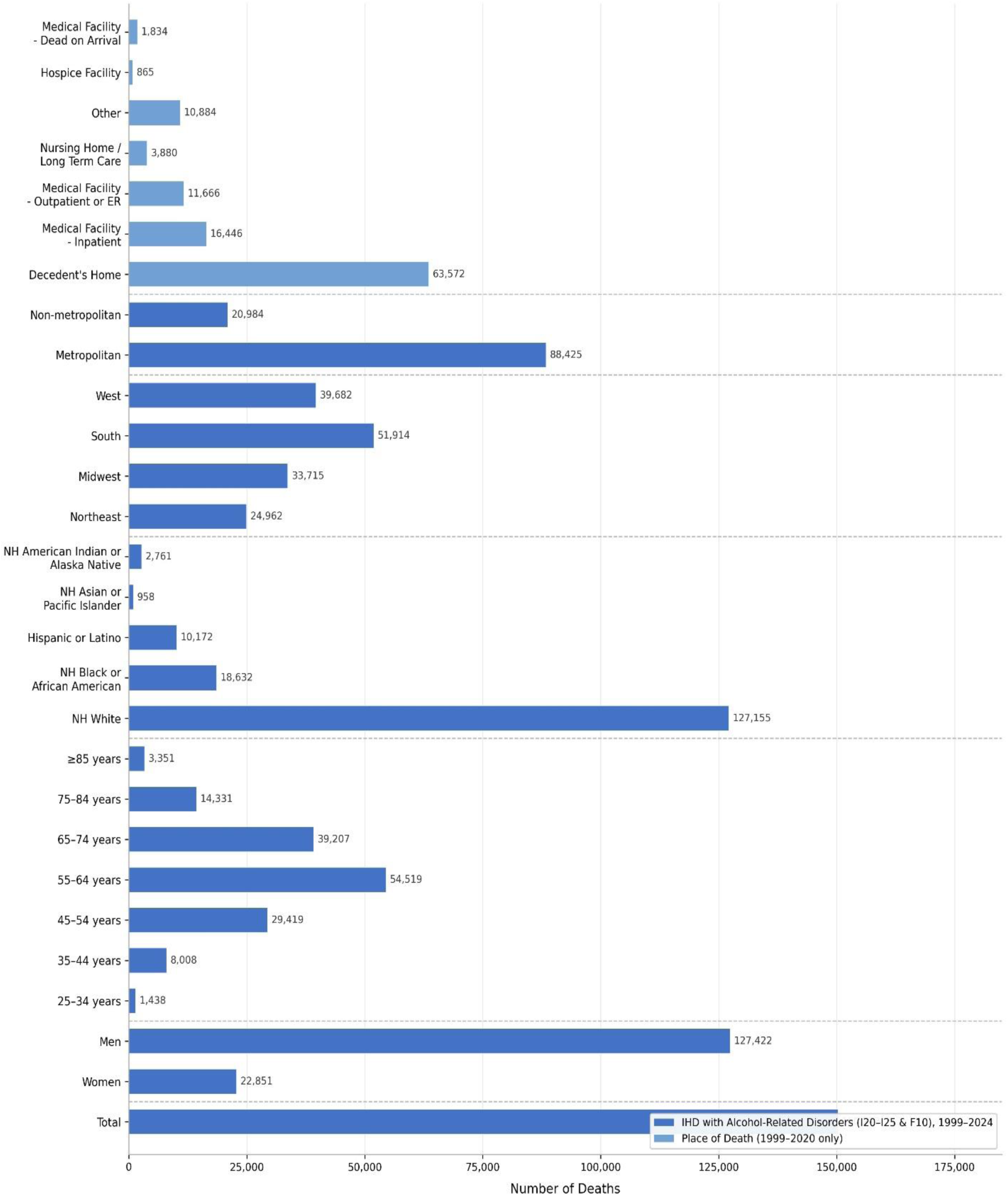
Total number of IHD+AUD-related deaths in the United States, 1999–2024, stratified by sex, age group, race/ethnicity, census region, urban-rural classification, and place of death. IHD indicates ischemic heart disease; AUD, alcohol use disorder; NH, non-Hispanic. Place of death data available for 1999–2020 only.

**Figure S2:**
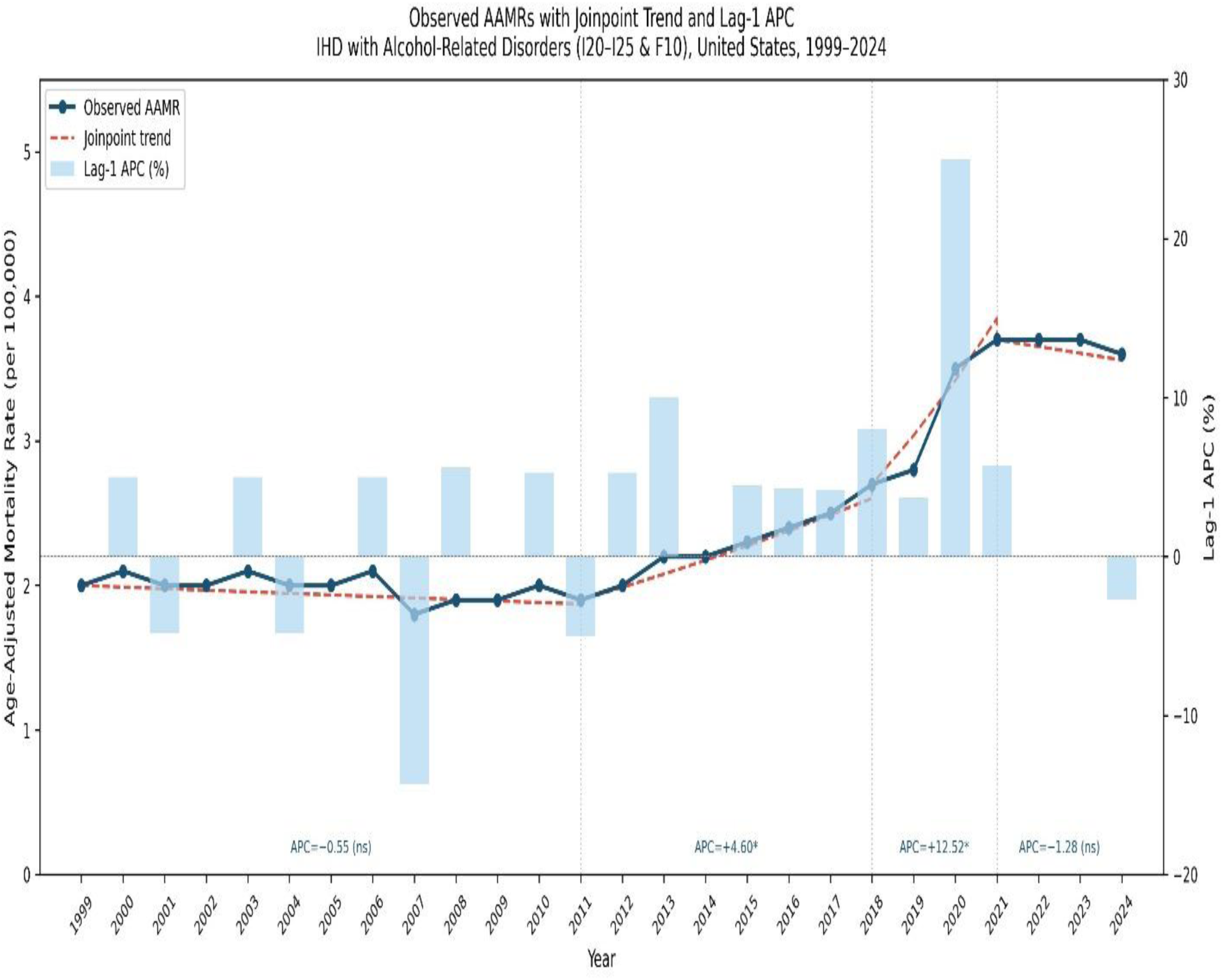
Overall age-adjusted mortality rates per 100,000 population and Joinpoint regression trend with Lag-1 annual percentage change for IHD with alcohol-related disorders (ICD-10 I20–I25 and F10) in the United States, 1999–2024. The Joinpoint-fitted trend is shown as a dashed red line overlaid on the observed AAMR. Lag-1 APC bars represent year-to-year percentage change in observed AAMR. Vertical dotted lines indicate statistically significant joinpoints. AAMR indicates age-adjusted mortality rate; APC, annual percentage change. *APC significantly different from zero at α=0.05. ns = not significant.

**Figure S3:**
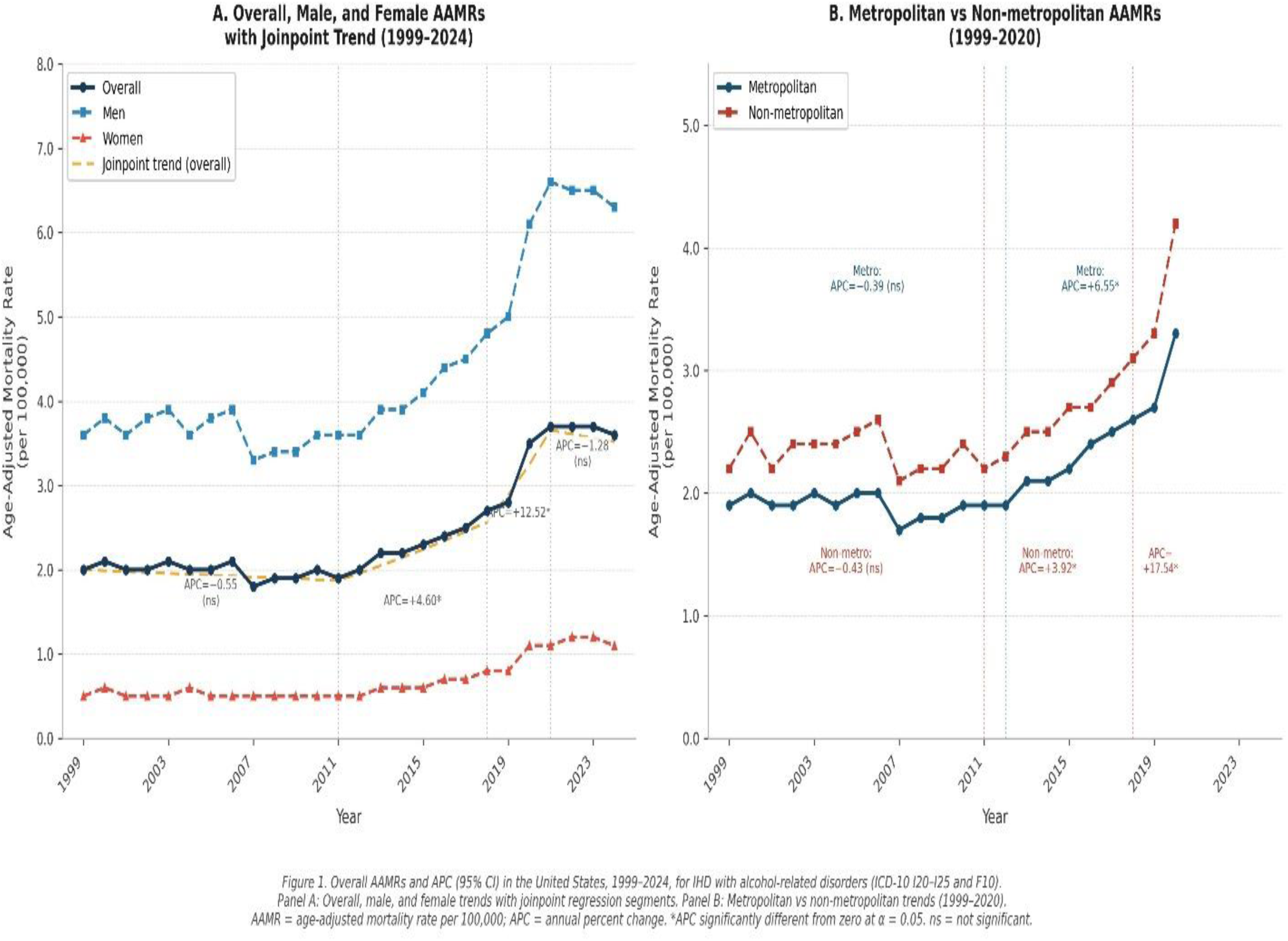
Two-panel figure: Panel A = overall, male, female AAMRs with joinpoint trends; Panel B = metropolitan vs non-metropolitan AAMRs

**Figure S4:**
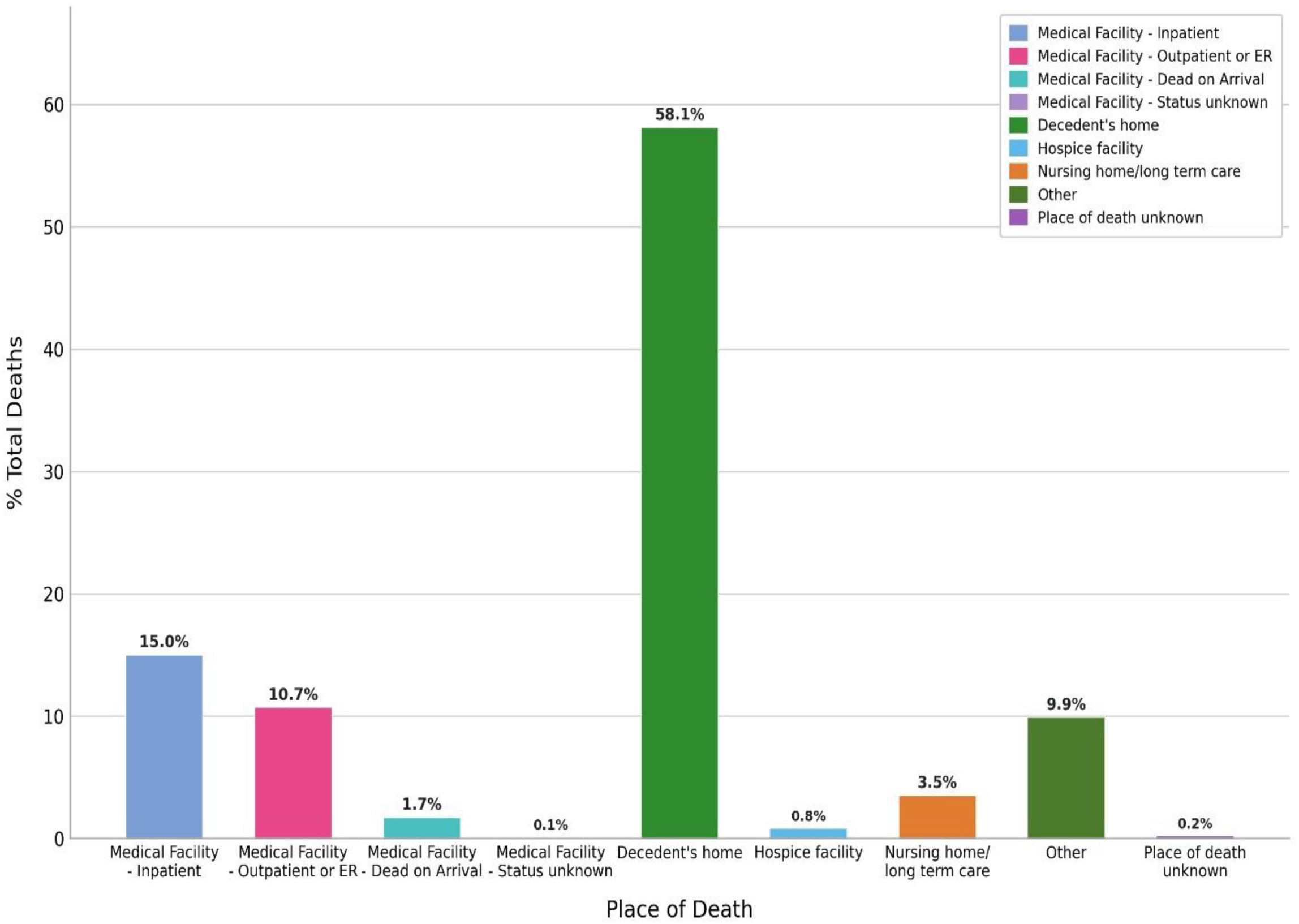
Distribution of place of death among IHD+AUD-related deaths in the United States, 1999–2020.

**Table S1.** Absolute Number of Ischemic Heart Disease with Alcohol-Related Disorders (ICD-10 codes I20–I25 and F10) Mortality Stratified by Sex and Race/Ethnicity in the United States, 1999–2024.

| Year | Overall | Females | Males | NH White | NH Black or African American | NH Asian or Pacific Islander | NH American Indian or Alaska Native | Hispanic or Latino |
| --- | --- | --- | --- | --- | --- | --- | --- | --- |
| 1999 | 3,448 | 687 | 2,761 | 2,709 | 458 | — | 107 | 163 |
| 2000 | 3,769 | 755 | 3,014 | 2,985 | 477 | — | 121 | 173 |
| 2001 | 3,633 | 694 | 2,939 | 2,871 | 479 | 22 | 107 | 153 |
| 2002 | 3,813 | 719 | 3,094 | 2,990 | 500 | 30 | 113 | 169 |
| 2003 | 3,985 | 744 | 3,241 | 3,135 | 484 | 35 | 115 | 198 |
| 2004 | 3,900 | 776 | 3,124 | 3,063 | 465 | 24 | 107 | 219 |
| 2005 | 4,087 | 733 | 3,354 | 3,176 | 526 | 33 | 121 | 214 |
| 2006 | 4,391 | 796 | 3,595 | 3,452 | 509 | 39 | 131 | 243 |
| 2007 | 3,778 | 680 | 3,098 | 2,978 | 445 | 31 | 114 | 197 |
| 2008 | 3,967 | 733 | 3,234 | 3,104 | 466 | 30 | 93 | 250 |
| 2009 | 4,081 | 731 | 3,350 | 3,187 | 481 | 33 | 113 | 245 |
| 2010 | 4,411 | 783 | 3,628 | 3,479 | 491 | 24 | 117 | 275 |
| 2011 | 4,503 | 799 | 3,704 | 3,567 | 508 | 24 | 103 | 278 |
| 2012 | 4,616 | 826 | 3,790 | 3,635 | 503 | 24 | 101 | 320 |
| 2013 | 5,224 | 993 | 4,231 | 4,143 | 570 | 37 | 120 | 328 |
| 2014 | 5,364 | 984 | 4,380 | 4,222 | 564 | 32 | 125 | 385 |
| 2015 | 5,724 | 1,018 | 4,706 | 4,496 | 631 | 43 | 138 | 391 |
| 2016 | 6,266 | 1,168 | 5,098 | 4,947 | 685 | 51 | 142 | 412 |
| 2017 | 6,560 | 1,175 | 5,385 | 5,145 | 688 | 53 | 143 | 490 |
| 2018 | 7,138 | 1,339 | 5,799 | 5,612 | 753 | 60 | 152 | 524 |
| 2019 | 7,440 | 1,358 | 6,082 | 5,775 | 789 | 58 | 160 | 611 |
| 2020 | 9,311 | 1,826 | 7,485 | 7,186 | 1,027 | 68 | 217 | 773 |
| 2021 | 10,213 | 2,027 | 8,186 | 7,814 | 1,144 | 85 | 255 | 870 |
| 2022 | 10,242 | 2,085 | 8,157 | 7,817 | 1,141 | 87 | 264 | 893 |
| 2023 | 10,294 | 2,080 | 8,214 | 7,803 | 1,148 | 86 | 291 | 918 |
| 2024 | 10,115 | 2,001 | 8,114 | 7,640 | 1,108 | 88 | 212 | 1,000 |
| **Total** | **150,273** | **22,851** | **127,422** | **127,155** | **18,632** | **958** | **2,761** | **10,172** |

Abbreviations: NH = Non-Hispanic. — indicates suppressed/unreliable data (n<20) per CDC WONDER criteria.

**Table S2.**
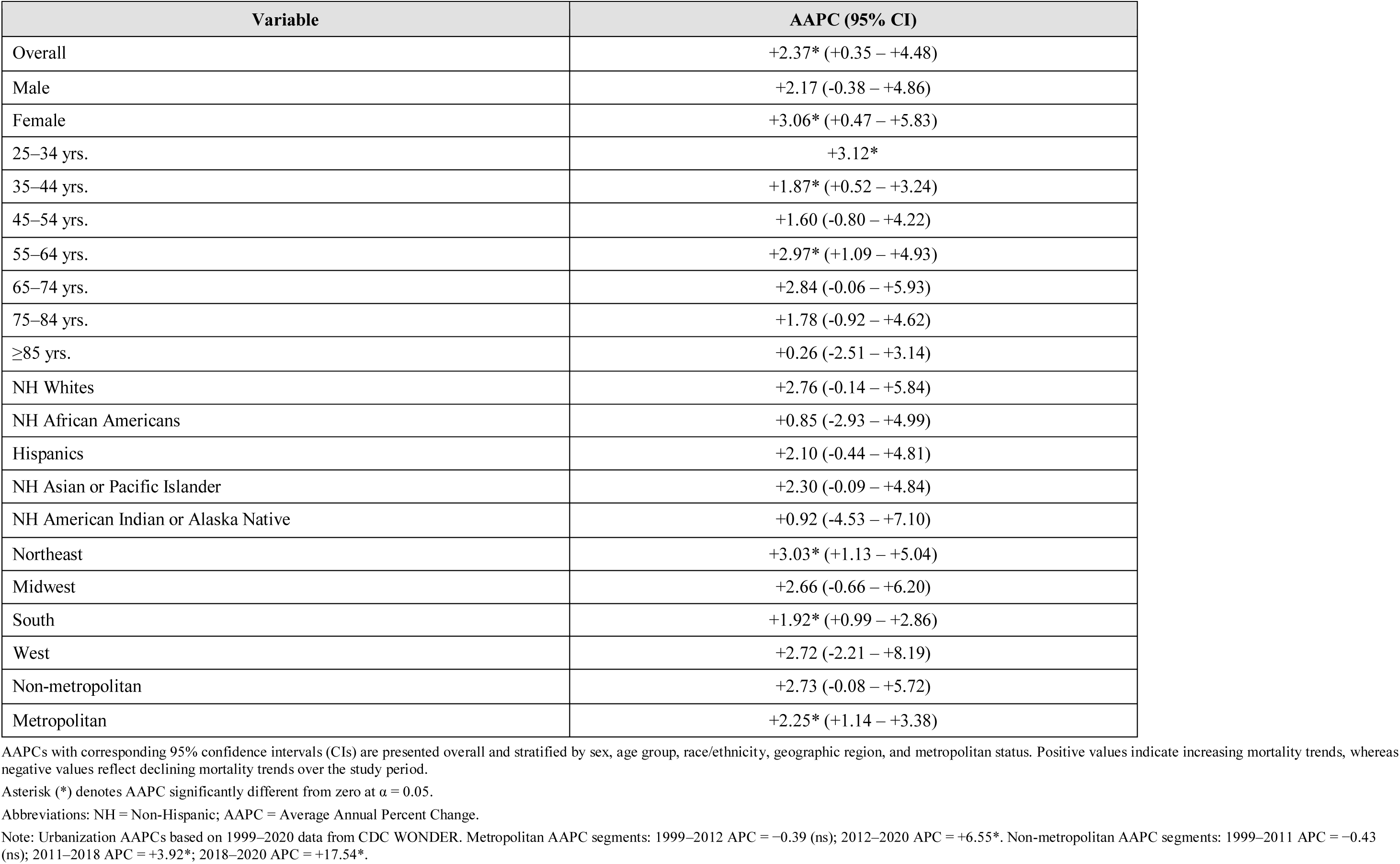
Average Annual Percent Change (AAPC) in Age-Adjusted Mortality Rates for Ischemic Heart Disease with Alcohol-Related Disorders (ICD-10 codes I20–I25 and F10), United States, 1999–2024.

**Table S3.**
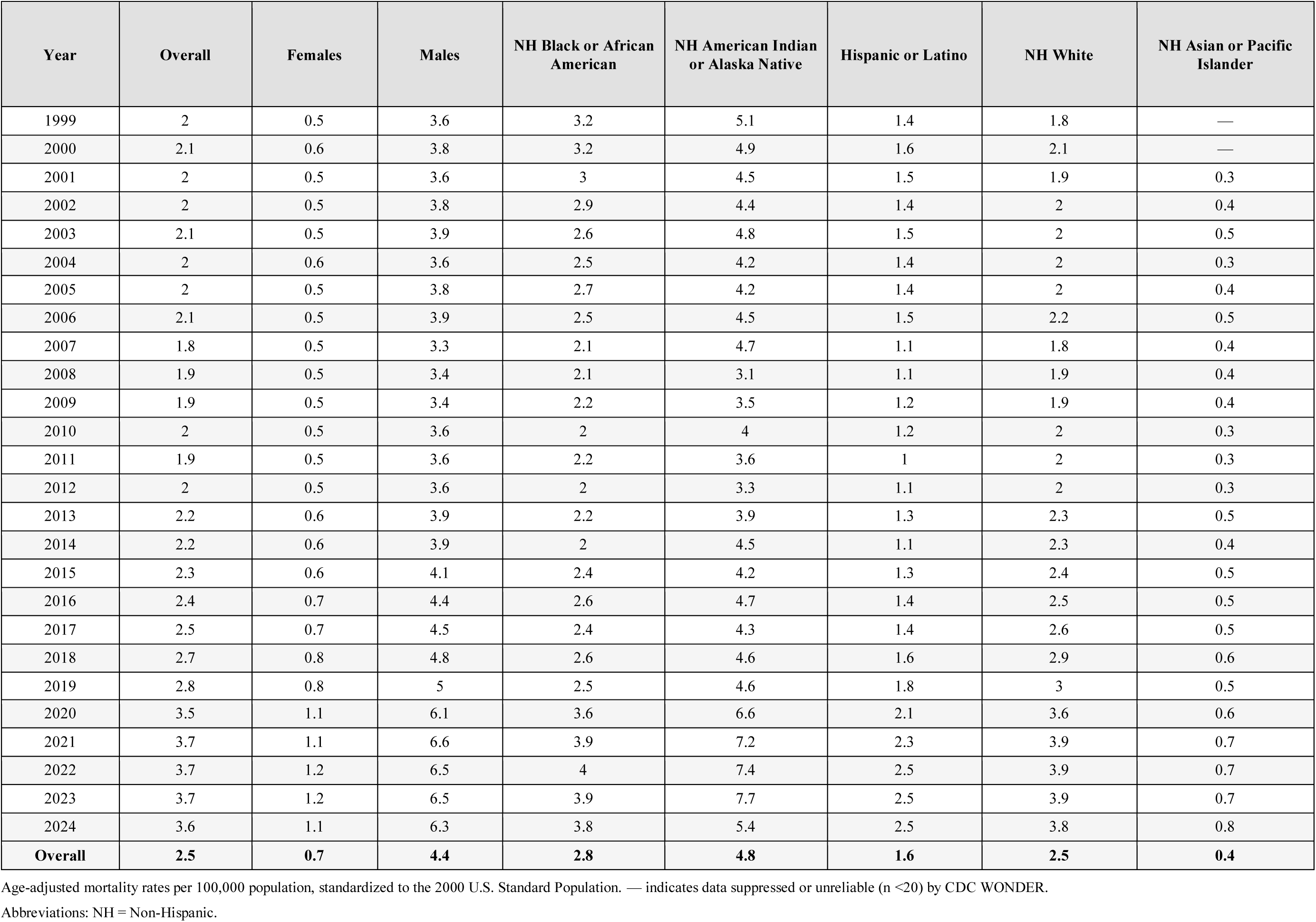
Ischemic Heart Disease with Alcohol-Related Disorders (ICD-10 codes I20–I25 and F10) Age-Adjusted Mortality Rates (per 100,000) Stratified by Overall Trend, Sex and Race/Ethnicity in the United States, 1999–2024.

**Table S4.** Ischemic Heart Disease with Alcohol-Related Disorders (ICD-10 codes I20–I25 and F10) Crude Mortality Rates (per 100,000) Stratified by Age Group in the United States, 1999–2024.

| Year | 25–34 years | 35–44 years | 45–54 years | 55–64 years | 65–74 years | 75–84 years | ≥85 years |
| --- | --- | --- | --- | --- | --- | --- | --- |
| 1999 | 0.1 | 0.6 | 2.1 | 3.9 | 4.9 | 3.8 | 2.9 |
| 2000 | 0.1 | 0.7 | 2.2 | 4.3 | 4.9 | 3.8 | 4.1 |
| 2001 | 0.1 | 0.7 | 2.3 | 4 | 4.7 | 3.4 | 2.2 |
| 2002 | 0.1 | 0.7 | 2.3 | 4.2 | 4.8 | 3.5 | 2.4 |
| 2003 | 0.1 | 0.7 | 2.5 | 4.4 | 4.7 | 3.3 | 2 |
| 2004 | 0.1 | 0.7 | 2.4 | 4.2 | 4.4 | 3.1 | 2.2 |
| 2005 | 0.1 | 0.6 | 2.5 | 4.3 | 4.4 | 3.4 | 2.2 |
| 2006 | 0.1 | 0.7 | 2.7 | 4.6 | 4.4 | 3.3 | 1.8 |
| 2007 | 0.1 | 0.5 | 2.3 | 3.9 | 4 | 2.8 | 1.9 |
| 2008 | 0.1 | 0.6 | 2.4 | 4 | 4 | 2.8 | 1.5 |
| 2009 | 0.1 | 0.5 | 2.3 | 4.1 | 4.3 | 2.9 | 1.5 |
| 2010 | 0.1 | 0.6 | 2.4 | 4.3 | 4.3 | 3.2 | 2.3 |
| 2011 | 0.1 | 0.5 | 2.6 | 4.4 | 4.2 | 2.8 | 1.8 |
| 2012 | 0.1 | 0.5 | 2.5 | 4.7 | 4 | 3.1 | 1.6 |
| 2013 | 0.1 | 0.5 | 2.7 | 5.2 | 4.8 | 3.1 | 1.8 |
| 2014 | 0.1 | 0.5 | 2.8 | 5.3 | 4.8 | 2.9 | 2 |
| 2015 | 0.1 | 0.5 | 2.7 | 5.5 | 5.2 | 3.5 | 1.6 |
| 2016 | 0.1 | 0.6 | 2.7 | 6.1 | 5.8 | 3.4 | 2.1 |
| 2017 | 0.1 | 0.6 | 2.7 | 6.4 | 5.8 | 3.7 | 2.1 |
| 2018 | 0.1 | 0.7 | 3 | 6.7 | 6.4 | 4.2 | 2 |
| 2019 | 0.2 | 0.8 | 2.8 | 6.8 | 6.8 | 4.3 | 2.3 |
| 2020 | 0.2 | 1.1 | 3.5 | 8.6 | 8.3 | 4.8 | 2.8 |
| 2021 | 0.2 | 1.2 | 3.7 | 9.1 | 9.3 | 5.3 | 3.2 |
| 2022 | 0.2 | 1.1 | 3.5 | 9.3 | 9.1 | 6 | 3.4 |
| 2023 | 0.2 | 1.3 | 3.4 | 9 | 9.3 | 5.7 | 3.6 |
| 2024 | 0.2 | 1.1 | 3.3 | 8.4 | 9.5 | 5.8 | 3 |
| **Overall** | **0.1** | **0.7** | **2.7** | **5.8** | **6** | **3.9** | **2.3** |

Crude mortality rates per 100,000 population. Age-adjusted rates are not presented at the age-group level as adjustment would be circular.
Abbreviations: IHD = Ischemic Heart Disease.

**Table S5.** Ischemic Heart Disease with Alcohol-Related Disorders (ICD-10 codes I20–I25 and F10) Age-Adjusted Mortality Rates (per 100,000) Stratified by Urban–Rural Classification in the United States, 1999–2020.

| **Year** | **Metropolitan** | **Non-metropolitan** |
| --- | --- | --- |
| 1999 | 1.9 (1.8–2.0) | 2.2 (2.0–2.3) |
| 2000 | 2.0 (1.9–2.1) | 2.5 (2.3–2.7) |
| 2001 | 1.9 (1.9–2.0) | 2.2 (2.1–2.4) |
| 2002 | 1.9 (1.9–2.0) | 2.4 (2.3–2.6) |
| 2003 | 2.0 (1.9–2.1) | 2.4 (2.3–2.6) |
| 2004 | 1.9 (1.8–2.0) | 2.4 (2.2–2.5) |
| 2005 | 2.0 (1.9–2.0) | 2.5 (2.3–2.6) |
| 2006 | 2.0 (2.0–2.1) | 2.6 (2.4–2.8) |
| 2007 | 1.7 (1.7–1.8) | 2.1 (1.9–2.2) |
| 2008 | 1.8 (1.7–1.8) | 2.2 (2.0–2.3) |
| 2009 | 1.8 (1.7–1.9) | 2.2 (2.1–2.4) |
| 2010 | 1.9 (1.8–1.9) | 2.4 (2.2–2.5) |
| 2011 | 1.9 (1.8–2.0) | 2.2 (2.1–2.4) |
| 2012 | 1.9 (1.8–2.0) | 2.3 (2.2–2.5) |
| 2013 | 2.1 (2.0–2.2) | 2.5 (2.3–2.6) |
| 2014 | 2.1 (2.0–2.2) | 2.5 (2.4–2.7) |
| 2015 | 2.2 (2.2–2.3) | 2.7 (2.5–2.8) |
| 2016 | 2.4 (2.3–2.4) | 2.7 (2.6–2.9) |
| 2017 | 2.5 (2.4–2.5) | 2.9 (2.7–3.0) |
| 2018 | 2.6 (2.6–2.7) | 3.1 (3.0–3.3) |
| 2019 | 2.7 (2.6–2.8) | 3.3 (3.2–3.5) |
| 2020 | 3.3 (3.3–3.4) | 4.2 (4.0–4.4) |
| **Overall** | **2.20 (2.20–2.20)** | **2.60 (2.40–2.70)** |

Age-adjusted mortality rates per 100,000 population, standardized to the 2000 U.S. Standard Population. 95% confidence intervals shown in parentheses.
Metropolitan includes Large Central Metro, Large Fringe Metro, Medium Metro, and Small Metro per 2013 NCHS urbanization classification. Non-metropolitan includes Micropolitan and NonCore categories.
Data limited to 1999–2020 due to CDC WONDER urbanization classification availability.

**Table S6.** State-Level Ischemic Heart Disease with Alcohol-Related Disorders (ICD-10 codes I20–I25 and F10) Age-Adjusted Mortality Rates per 100,000 People in the United States, 1999–2020.

| Rank | Percentile | State | AAMR per 100,000 individuals |
| --- | --- | --- | --- |
| 1 | 0 | Alabama | 0.9 |
| 2 | 2 | Hawaii | 1.4 |
| 3 | 4 | Nebraska | 1.4 |
| 4 | 6 | Georgia | 1.5 |
| 5 | 8 | New Jersey | 1.5 |
| 6 | 10 | Illinois | 1.5 |
| 7 | 12 | Pennsylvania | 1.5 |
| 8 | 14 | Missouri | 1.6 |
| 9 | 16 | Virginia | 1.6 |
| 10 | 18 | Louisiana | 1.7 |
| 11 | 20 | Connecticut | 1.8 |
| 12 | 22 | Texas | 1.8 |
| 13 | 24 | Utah | 1.8 |
| 14 | 26 | Indiana | 2 |
| 15 | 28 | Massachusetts | 2 |
| 16 | 30 | Michigan | 2.1 |
| 17 | 32 | Florida | 2.1 |
| 18 | 34 | Maine | 2.2 |
| 19 | 36 | New York | 2.2 |
| 20 | 38 | South Carolina | 2.3 |
| 21 | 40 | Mississippi | 2.3 |
| 22 | 42 | California | 2.3 |
| 23 | 44 | Kentucky | 2.3 |
| 24 | 46 | Maryland | 2.4 |
| 25 | 48 | Tennessee | 2.4 |
| 26 | 50 | Ohio | 2.4 |
| 27 | 52 | Arkansas | 2.5 |
| 28 | 54 | Idaho | 2.6 |
| 29 | 56 | New Hampshire | 2.7 |
| 30 | 58 | Kansas | 2.7 |
| 31 | 60 | Arizona | 2.8 |
| 32 | 62 | North Carolina | 2.9 |
| 33 | 64 | Oregon | 2.9 |
| 34 | 66 | Washington | 2.9 |
| 35 | 68 | Iowa | 3.1 |
| 36 | 70 | West Virginia | 3.1 |
| 37 | 72 | Minnesota | 3.1 |
| 38 | 74 | South Dakota | 3.2 |
| 39 | 76 | Delaware | 3.3 |
| 40 | 78 | Colorado | 3.4 |
| 41 | 80 | Oklahoma | 3.4 |
| 42 | 82 | Wisconsin | 3.6 |
| 43 | 84 | Rhode Island | 3.6 |
| 44 | 86 | Wyoming | 3.7 |
| 45 | 88 | Nevada | 4.1 |
| 46 | 90 | New Mexico | 4.1 |
| 47 | 92 | Montana | 4.2 |
| 48 | 94 | District of Columbia | 4.5 |
| 49 | 96 | Alaska | 4.5 |
| 50 | 98 | North Dakota | 4.9 |
| 51 | 100 | Vermont | 5.2 |
|  |  | **U.S.A** | **2.5** |

States ranked in ascending order of age-adjusted mortality rate. Percentile rank calculated across 51 jurisdictions (50 states + District of Columbia).
Abbreviations: AAMR = Age-Adjusted Mortality Rate; U.S.A = United States of America.

**Table S7.**
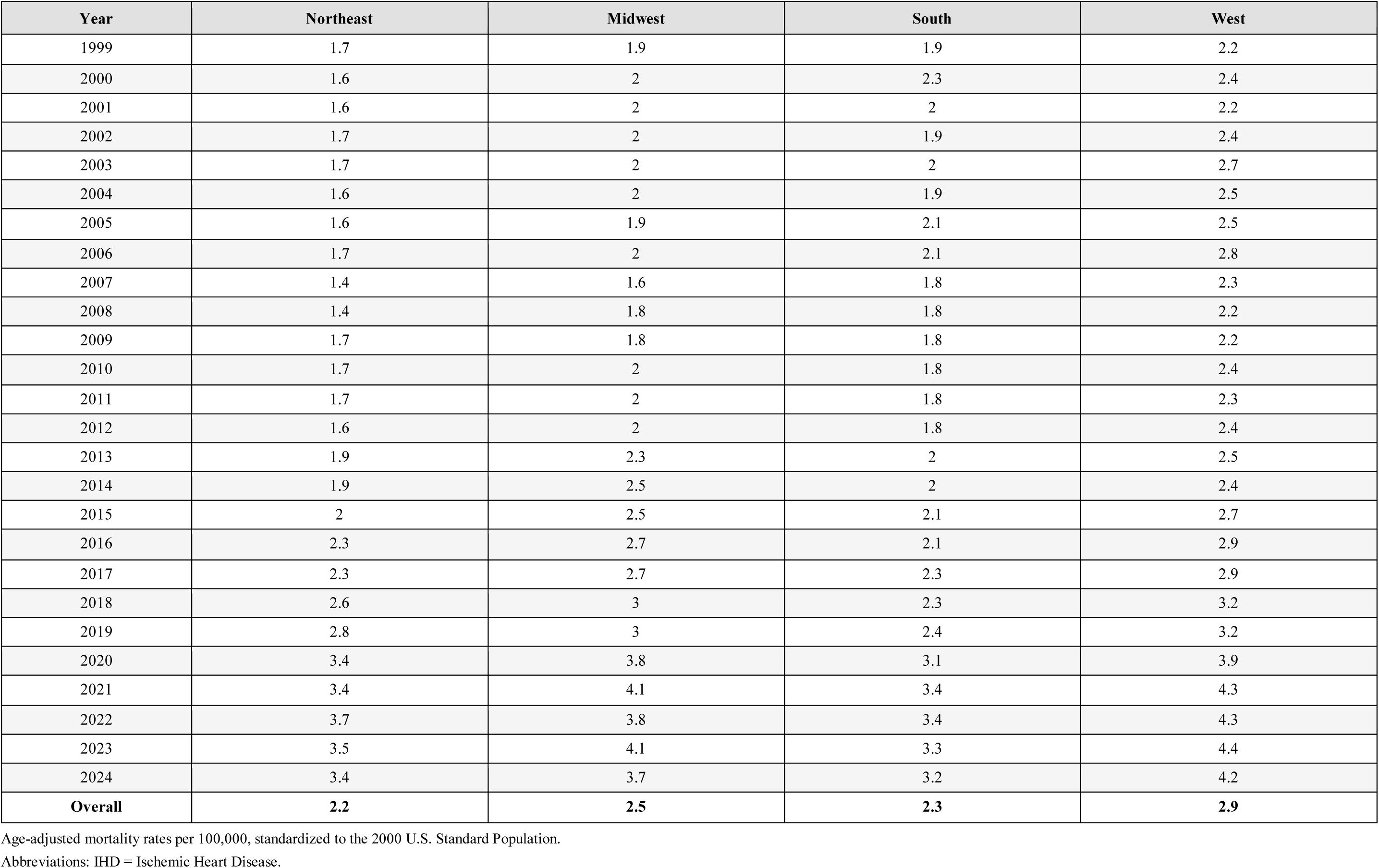
Ischemic Heart Disease with Alcohol-Related Disorders (ICD-10 codes I20–I25 and F10) Age-Adjusted Mortality Rates (per 100,000) Stratified by Census Region in the United States, 1999–2024.

